# The mathematics of erythema: Development of machine learning models for artificial intelligence assisted measurement and severity scoring of radiation induced dermatitis

**DOI:** 10.1101/2021.09.24.21264011

**Authors:** Rahul Ranjan, Richard Partl, Ricarda Erhart, Nithin Kurup, Harald Schnidar

## Abstract

Although significant advancements in computer-aided diagnostics using artificial intelligence (AI) have been made, to date, no viable method for radiation-induced skin reaction (RISR) analysis and classification is available. The objective of this single-center study was to develop machine learning and deep learning approaches using deep convolutional neural networks (CNNs) for automatic classification of RISRs according to the Common Terminology Criteria for Adverse Events (CTCAE) grading system. Scarletred^®^ Vision, a novel and state-of-the-art digital skin imaging method capable of remote monitoring and objective assessment of acute RISRs was used to convert 2D digital skin images using the CIELAB color space and conduct SEV* measurements. A set of different machine learning and deep convolutional neural network-based algorithms has been explored for the automatic classification of RISRs. A total of 2263 distinct images from 209 patients were analyzed for training and testing the machine learning and CNN algorithms. For a 2-class problem of healthy skin (grade 0) versus erythema (grade ≥ 1), all machine learning models produced an accuracy of above 70%, and the sensitivity and specificity of erythema recognition were 67-72% and 72-83%, respectively. The CNN produced a test accuracy of 74%, sensitivity of 66%, and specificity of 83% for predicting healthy and erythema cases. For the severity grade prediction of a 3-class problem (grade 0 versus 1 versus 2), the overall test accuracy was 60-67%, and the sensitivities were 56-82%, 35-59%, and 65-72%, respectively. For estimating the severity grade of each class, the CNN obtained an accuracy of 73%, 66%, and 82%, respectively. Ensemble learning combines several individual predictions to obtain a better generalization performance. Furthermore, we exploited ensemble learning by deploying a CNN model as a meta-learner. The ensemble CNN based on bagging and majority voting shows an accuracy, sensitivity and specificity of 87%, 90%, and 82% for a 2-class problem, respectively. For a 3-class problem, the ensemble CNN shows an overall accuracy of 66%, while for each grade (0, 1, and 2) accuracies were 76%, 69%, and 87%, sensitivities were 70%, 57%, and 71%, and specificities were 78%, 75%, and 95%, respectively. This study is the first to focus on erythema in radiation-dermatitis and produces benchmark results using machine learning models. The outcome of this study validates that the proposed system can act as a pre-screening and decision support tool for oncologists or patients to provide fast, reliable, and efficient assessment of erythema grading.

## 1. Introduction

Radiotherapy is the primary treatment for several types of malignant tumors and has become a central and effective component of modern oncologic strategies. One of the most common side effects of therapeutic ionizing radiation is an acute radiation-induced skin reaction (RISR), which is characterized as structural tissue damage, nonspecific inflammation, and the release of free radicals, interacting and promoting each other [1]. Depending on the total dose applied, fractionation scheme, volume of irradiated skin, and the patient’s individual radiosensitivity, acute changes to the skin become visible and often lead to pain, discomfort, itching, and a diminished esthetic appearance, all of which affect the patient’s quality of life and the progress of the cancer treatment [2, 3, 4]. In clinical practice, acute RISRs are still evaluated using subjective visual classifications such as those provided by the Common Terminology Criteria for Adverse Events (CTCAE) and the Radiation Therapy Oncology Group/European Organization for Research and Treatment of Cancer (RTOG/EORTC) [5, 6]. According to the degree of severity, the morphological changes are divided into distinct grades, starting with grade 1, characterized by faint erythema or dry desquamation, and ending with skin necrosis and ulceration of the dermis in grade 4 reactions (Table 1).

**Table 1:** Injury, poisoning and procedural complications: Different CTCAE grades and their clinical description published by the U.S. DoHSS Definition: A reaction occurring as a result of exposure to biologically effective levels of ionizing radiation. Navigational Note: Synonym: Radiation induced skin toxicities (CTCAE v4.03)

| CTCAE Terms: | Grade 1 | Grade 2 | Grade 3 | Grade 4 | Grade 5 |
| --- | --- | --- | --- | --- | --- |
| Dermatitis radiation | Faint erythema or dry desquamation | Moderate to brisk erythema; patchy moist desquamation, mostly confined to skin folds and creases; moderate edema | Moist desquamation in areas other than skin folds and creases; bleeding induced by minor trauma or abrasion | Life threatening consequences; skin necrosis or ulceration of full thickness dermis; spontaneous bleeding from involved site; skin graft indicated | Death |

However, a purely visual inspection is prone to a considerable risk of interobserver variability [7]. To overcome these shortcomings, efforts have been made to develop alternative approaches for erythema assessment. An increasing number of data have demonstrated that methods objectively measuring the skin color or dermal oxygenation of hemoglobin can provide additional information [8, 9, 10, 11, 12]. Spectra-based methods illuminate the patient’s skin with a certain wavelength or whole spectrum and measure the reflected light. The instruments are comparably inexpensive and yield reliable results; however, they have to be applied to patients irradiated with skin, which can be very painful. Furthermore, the area of the skin that can be measured is extremely small. Therefore, it is sometimes necessary to perform multiple measurements to obtain reproducible results; however, this makes the data collection and analysis process time intensive, and the method is thus ineligible for everyday clinical use. Another approach is applying an image-based method that enables an image to be taken under standardized light conditions. To obtain standardized images, a large amount of expensive equipment and expertise is required. In addition, this method requires no contact with the skin and large areas can be evaluated [13]. Hyperspectral imaging combines the advantages of both methods by illuminating the skin and measuring the reflected light at a distance. This new method has yielded the most promising results [14, 15]. However, a lack of standardization remains, and a time-consuming preparation as well as the high cost of a manual analysis have limited the application of these new methods in routine clinical practice.

In one of our previous studies, we introduced Scarletred^®^ Vision, a novel and state-of-the-art digital skin imaging method capable of remote monitoring and objective assessment of acute RISRs [16]. A normalization patch was attached to the region of interest and a color-calibrated picture was taken with a smartphone app. After marking the area of interest, the images were automatically transformed into the three-dimensional CIELAB color space defined by the Commission International d’Eclairage (CIE). The three coordinates are the *L*\* parameter, which stands for the lightness; the *a*\* parameter, which reflects the green/red axis; and the *b*\* parameter, which reflects the blue/ yellow axis. This patented medical device overcomes the pitfalls of previous described image- and spectra-based methods by introducing a standardized erythema value (*SEV* *) derived from the algorithm (*L*\*max - *L*\*) x +*a*\* in combination with a color normalization sticker [17]. From a practical standpoint, we believe that there is a high need for further automation of the image-analysis method to find its applicability in daily clinical practice. Overcoming this last hurdle requires further developing and integrating novel state-of-the-art machine learning (ML) methods.

In recent years, artificial intelligence (AI) has increasingly found its way into clinical research as well as in medical routines. AI algorithms have already been successfully used for diagnostic [18, 19, 20] and predictive tasks [21] and deep learning neural networks have led to state-of-the-art results for image classification, object detection, and image segmentation in a broad field of medical applications. Several studies have demonstrated that AI algorithms are even superior to a clinician performance [22]. By contrast, the use of AI approaches for the classification of acute RISRs has been relatively less studied. In this study, we aimed at developing ML models and a novel deep convolutional neural network (CNN) based architecture to automatically analyze and classify the RISR severity according to the CTCAE grading system (Figure 1).

**Figure 1:**
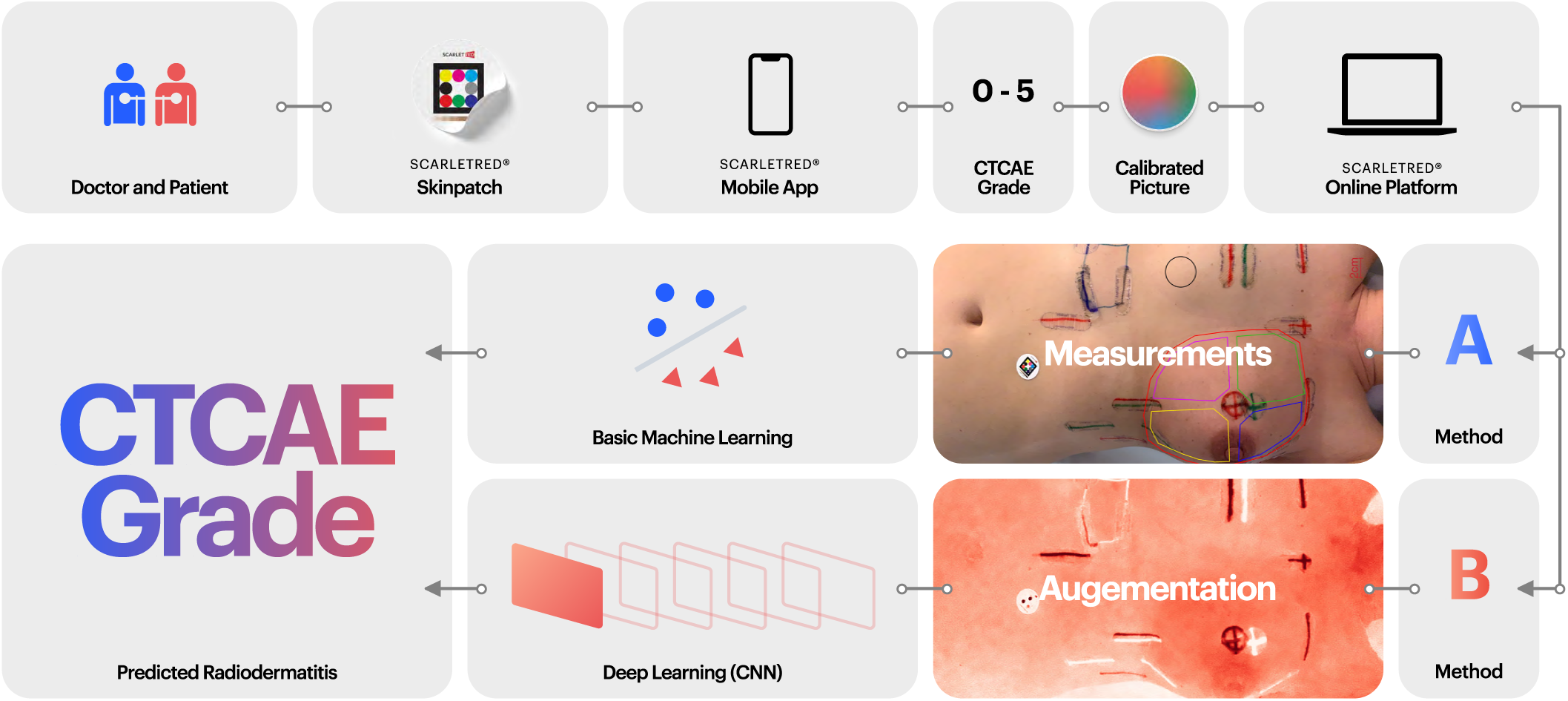
Workflow from clinical image to machine learning and deep learning-based CTCAE grade classification. A skin reaction is visually assessed according to CTCAE criteria. Using the Scarletred^®^Vision app and a calibration sticker, an image is taken and uploaded to the online platform. The images are then analyzed by measuring L*, +a*, +b*, and SEV * values in defined areas of interest and then augmented. They are then used in machine learning and deep learning respectively. To train both a CNN and standard machine learning models, CTCAE grades from a visual assessment are used and the trained algorithms are applied to automatically predict the CTCAE grade for new images or measurements.

## 2. Materials and methods

### 2.1. Study design and patient demographics

In this prospective clinical single-center study, 209 consecutive patients with histologically confirmed cancer and treated with radiotherapy between January and July 2019 at a tertiary academic center were included. The median age in years was 67.8 (range of 36.3-86.2), 65.9 (mean), and ±12 (SD). In this study, 25.4% had fair skin and blue eyes (Fitzpatrick type I-II), 67.9% had darker olive to light brown skin (Fitzpatrick type III), 6.2% had light brown skin (Fitzpatrick type IV), and 0.5% had dark brown skin (Fitzpatrick type V) (Table 2). The study complied with the Declaration of Helsinki and was conducted according to the national law. The study protocol was approved by the local ethical committee (approval number, EK 30-023 ex 17/18 from 06 March 2018). All patients provided informed consent.

**Table 2:**
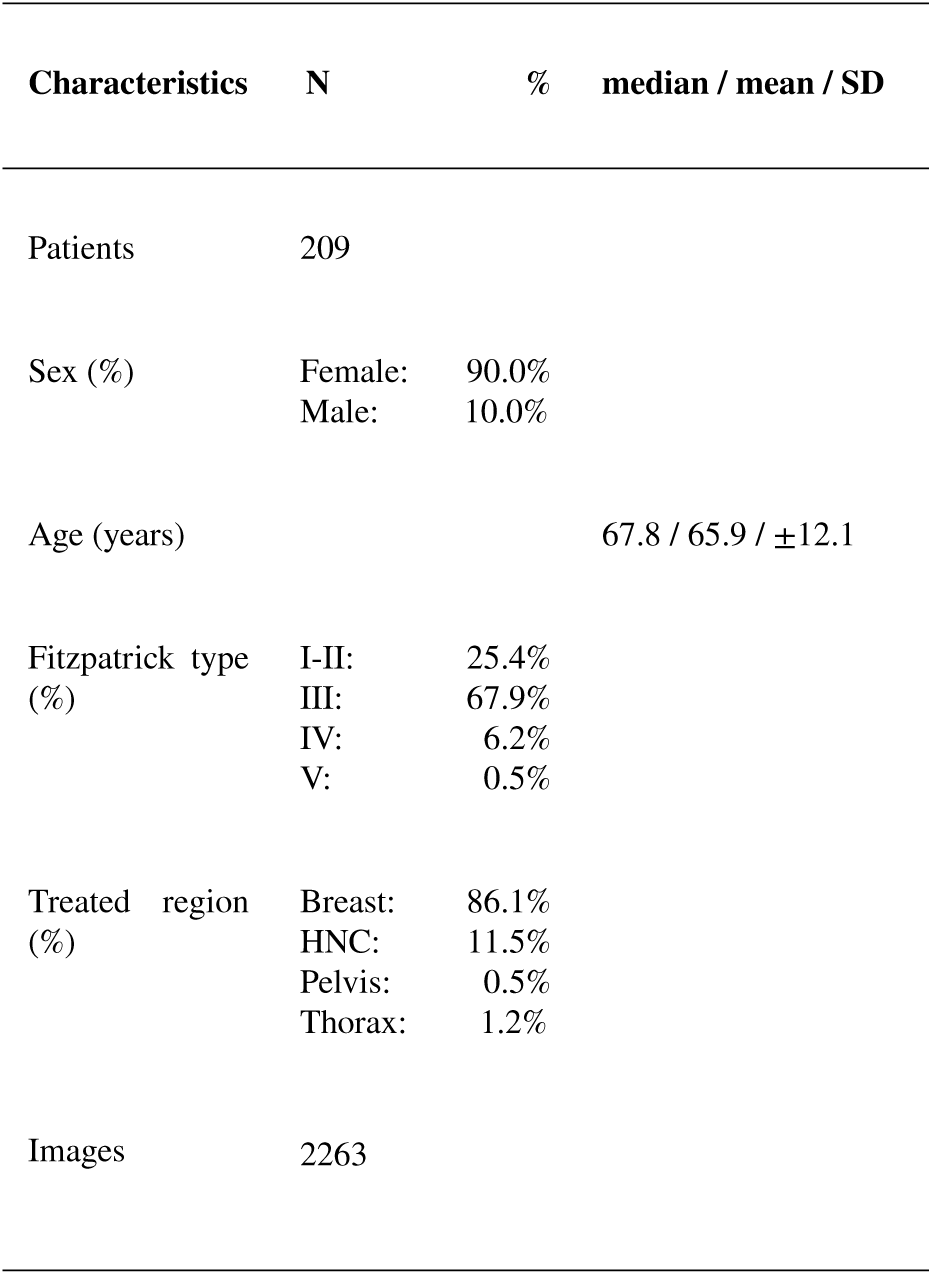
Patient characteristics (N = 209)

### 2.2. Radiotherapy setup

All patients were irradiated in a clinical routine setup using Clinac^®^ iX and Novalis Tx system linear accelerators. Depending on the tumor stage, localization, and performance status, the radiation method is either three-dimensional conformal radiotherapy or intensity-modulated radiotherapy (IMRT), including volumetric modulated arc therapy (VMAT) with normal or hypofractionated photon beams. According to the study protocol, RISR was scored daily by a board-certified radiation oncologist, based on the EORTC/RTOG-CTCAE v4.03 classification system beginning at baseline before the start of radiotherapy.

### 2.3. Image analysis

Skin imaging conditions play an important role in the success of remote classification. In total, 2263 distinct images of the irradiated area were taken before the treatment and at regular intervals during the treatment by using a CE certified medical device, i.e., Scarletred^®^Vision. To reflect a more realistic clinical scenario, the study design foresaw variable acquisition conditions in terms of illumination, viewpoint, and background. Before taking an image, a Scarletred^®^ Skin patch was applied to the healthy skin of the patients next to the irradiated area. The Scarletred^®^ Mobile App automatically recognized the Scarletred^®^ Skin Patch and enabled standardization of exposure, colors, imaging distance, and angle. The image was uploaded to the Scarletred^®^ Online Platform, and the total irradiated area was marked for a subsequent image analysis. In breast cancer, the analysis area was further subdivided into four quadrants to exclude irradiation markings (Figure 1). A single reference area was drawn around the patch to normalize the healthy skin and individual skin tone of the person. For each pixel inside a drawn area, the *L*\*, +*a*\* (posA), and +*b*\* (posB) coordinates of the CIELAB-color space and the standardized erythema value (*SEV* *) were determined. The means of *L*\*, +*a*\*, +*b*\*, and *SEV* * for every area were saved in the database.

In addition to the “mean” values, the difference of these values over time was calculated, resulting in the “meandt” value for every analyzed parameter. Further, the difference over time was calculated for the reference area. This value was then subtracted from the “meandt” value of the area of interest in the same picture. The resulting “meandt-refdt” value was saved in the database and used in the following analysis. The images were scored according to the CTCAE v4.03 criteria by a board-certified radiation oncologist. Continuous parameters were expressed as the mean, standard deviation, median, interquartile range, minimum, and maximum and visualized using histograms, boxplots, and QQ-plots. Categorical data were presented as absolute and relative frequencies, and bar charts were used for visualization.

Boxplots were used to visualize the differences in the measurements between images scored with different CTCAE grades based on a visual assessment. Furthermore, statistical hypothesis tests were applied to our biostatistics tool to determine if the groups were significantly different. Since the underlying distribution of the data often did not follow a normal distribution (Supplementary Figure 14), we used a non-parametric test to compare the different groups. To avoid overpowered testing, which is the generation of significant results from small differences between groups owing to the large sample size, we chose a random sample of 35 data points per group. The Kruskal-Wallis test was applied to compare the measurements (meandt-refdt) of multiple groups at a significance level of 5%.

### 2.4. Signal intensity mapping

The calculated signal intensity map was extracted from the original images used as gray scale value images, and subsequently, the high dynamic pseudo-gray scale images were transformed into pseudo-colored images to optimize for human perception. Each signal intensity value was mapped to a color according to our developed color maps, which were designed and tailored to the underlying signal (Figure 2). Each map defines a color gradient over the minimum to maximum signal range, which enables the visualization of more details and potential signal saturation.

**Figure 2:**
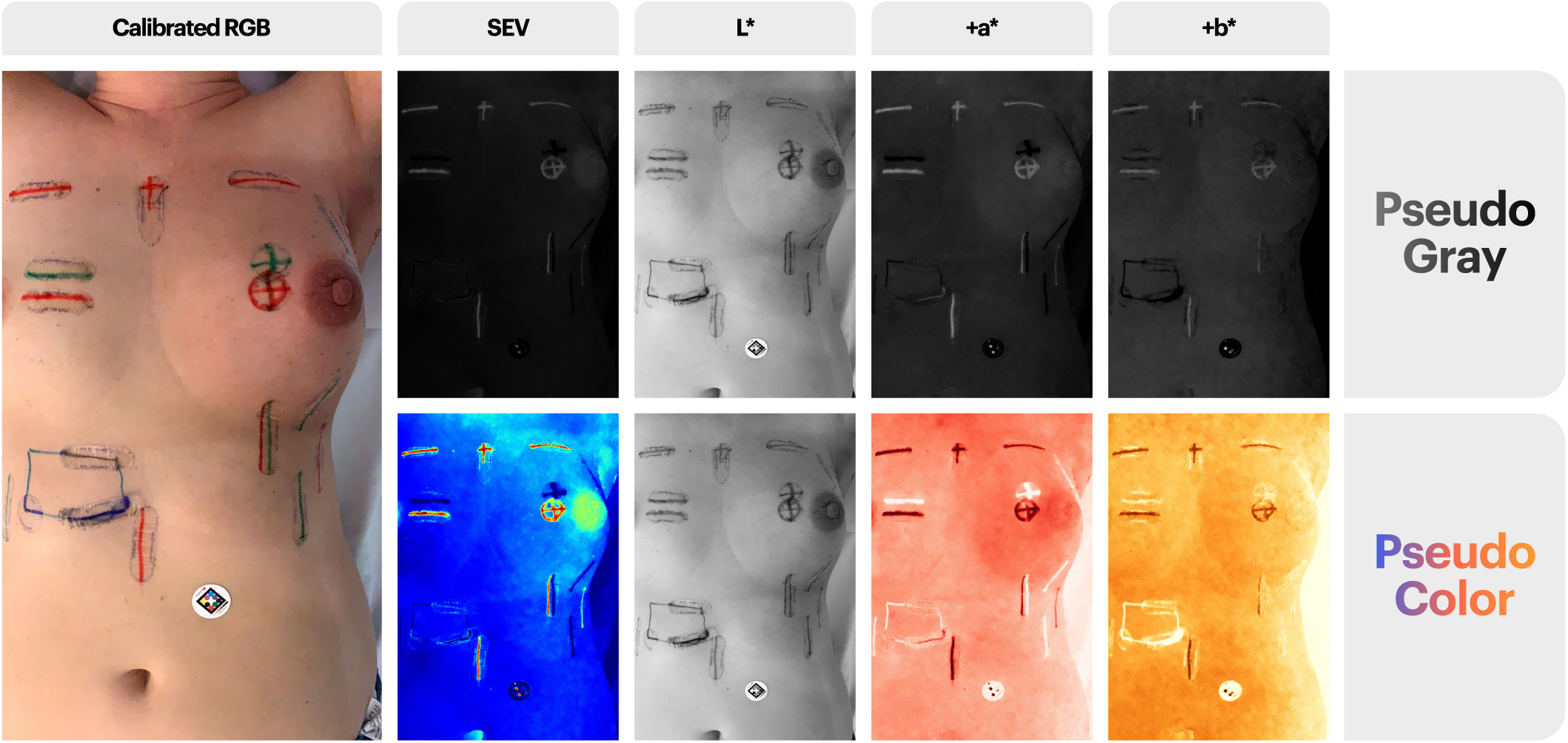
Calibrated RGB original image is shown on the left. Images were signal augmented and analysed by measuring the SEV *, L*, +a* and +b* values in pseudo gray scale in defined areas of interest, as shown in the top row. The signal intensity map was generated using pseudo-gray scale value images and by mapping each signal value to a pseudo color according to our developed color maps, which are designed and tailored to the underlying signals. In this study, the pseudo-gray scale images were used for training the CNN algorithms. In the first and second rows, the pseudo-gray scale and pseudo-colored signal intensity mapped images are in the following order: SEV *, L*, +a* (posA) and +b* (posB)

### 2.5. Machine learning

To deduce the erythema grade from *L*\*, +*a*\*, +*b*\*, and *SEV* * (meandt-refdt) measurements, machine learning models were built, including multinomial logistic regression, decision trees, random forest, and support vector machines. Because *L*\* and +*a*\* show a high correlation to *SEV* *, we used either the former two or the latter parameter as a feature. In addition, we also used +*b*\* as the potential feature. To build and evaluate the models, the dataset was split randomly into two groups, 75% were assigned to the training set and the remaining 25% were assigned to the test set. To compare the different machine learning models, three efficacy parameters were used (Table 3). The overall accuracy indicates what percentage of cases the machine learning and visual assessment resulted in the same grade. The sensitivity (true positive rate) was calculated as the number of true positive results divided by the sum of all the true positive and false negative results, whereas the specificity (true negative rate) was calculated as the number of true negative results divided by the sum of all true negative and false positive results. Furthermore, the model with the highest accuracy was visualized using a confusion matrix and ROC curves. ROC curves then plot the false positive rate on the x-axis against the true positive rate on the y-axis for every grade. The false positive rate (1-specificity) is all false positive instances divided by all true negative and false positive instances.

**Table 3:** Overview on the meaning of true positive, false positive, false negative, true negative, false positive rate, true positive rate, sensitivity, specificity, and accuracy. It is exemplary for classification into CTCAE grade, which can either be positive or negative (any other grade) in a visual assessment and random forest.

| Classification in CTCAE-grade | Positive outcome: visual assessment | Negative outcome: visual assessment |
| --- | --- | --- |
| Positive outcome: ML models | True positive (TP) | False positive (FP) |
| Negative outcome: ML models | False negative (FN) | True negative (TN) |

### 2.6. CNN classification

In this task, we implemented a CNN algorithm to distinguish between healthy and erythematous classes along with estimating the severity grade. The input dataset consisted of pseudo gray images resulting from *L*\*, +*a*\*, +*b*\*, and *SEV* * signal mapping. A total of 75% of the dataset was used for training, whereas the remaining 25% was used for testing.

Once the images were processed and collected, we implemented a series of pre-processing techniques, including data preparation and augmentation. For data augmentation, the input images were rescaled (1./255), rotated 15–45 degrees, shuffled, batched, and resized to a pixel resolution of 128 × 128. Figures 3 and 4, provide a visual representation of the total number of images for training CNNs; a closer and representative look on single images is provided in Figure 2). The images in the figures show randomness since the input dataset has been shuffled to prevent the model from overfitting and to generalise better on test dataset. The figures contain images from different color-spaces (*SEV* *, *L*\*, +*a*\*, and +*b*\*). Data augmentation is one of the approaches used to reduce the chances of overfitting; however, it is insufficient because the augmented data are still highly correlated (we therefore implemented a dropout and regularization to prevent an overfitting). Multiple approaches have been implemented to prevent the model from overfitting, including dropouts, early stopping, and regularization. Further, we implemented ensemble learning to reduce the effect of the variance in the input data. According to Shen et al. the neural networks (NNs) are the main building blocks of the deep learning architecture [23], which has three actions: receiving an input, processing information and generating an output. A neural network consists of an input layer, hidden layers, and an output layer. The feed-forward neural network is the simplest form of a neuronal network (Figure 5). It comprises weights, activation functions, and biases.

**Figure 3:**
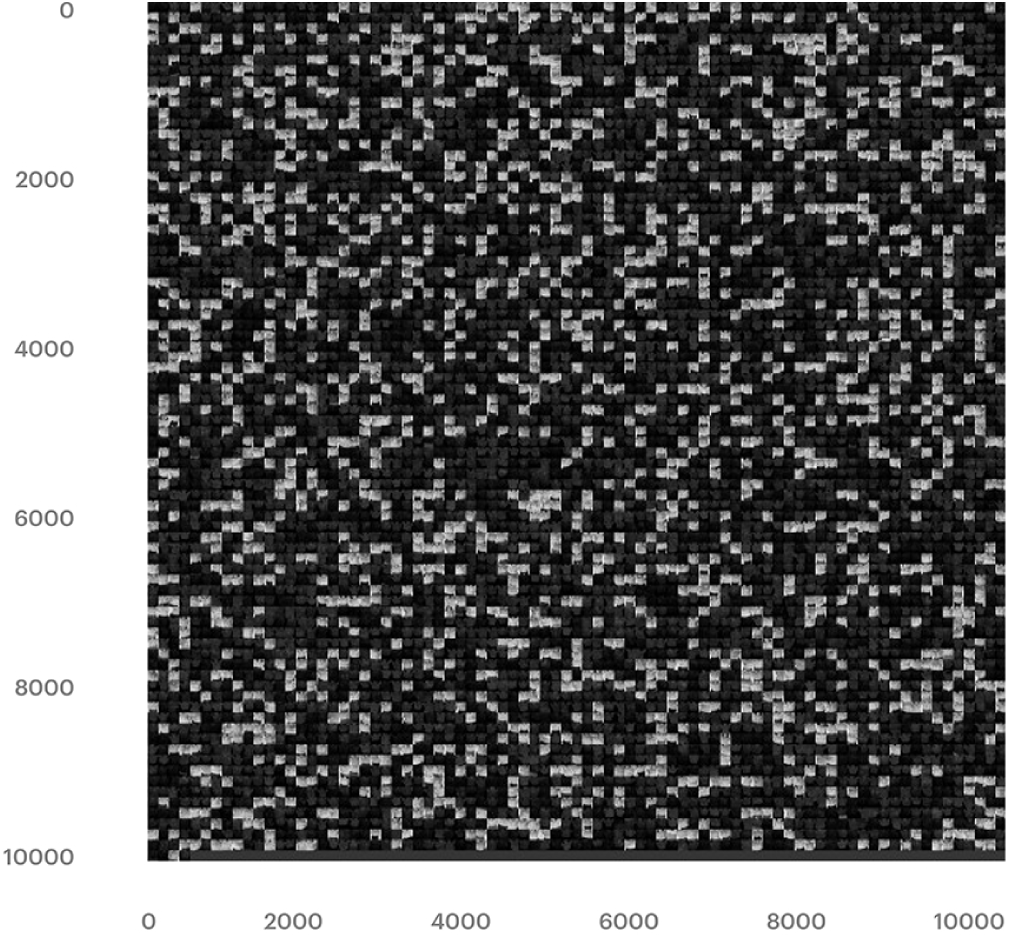
Input data (pseudo-gray) used for training the CNN architecture. The images are from L*, +a*, +b*, and SEV *. During the image augmentation technique, images were re-scaled by 1./255, rotated, batched, shuffled, and resized images.

**Figure 4:**
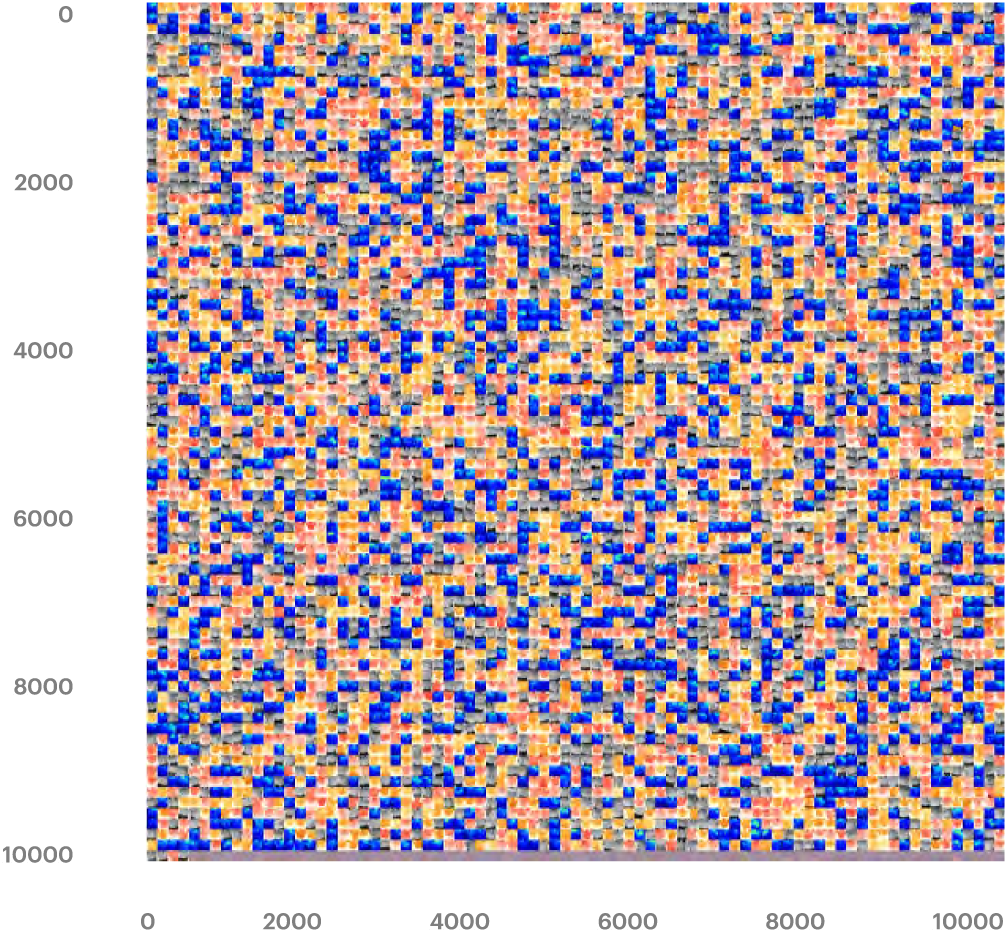
Pseudo-color input data for training a CNN. These images belong to the L*, +a*, +b*, and SEV * color-space. During the image augmentation technique, images were re-scaled by 1./255, rotated, batched, shuffled, and resized.

**Figure 5:**
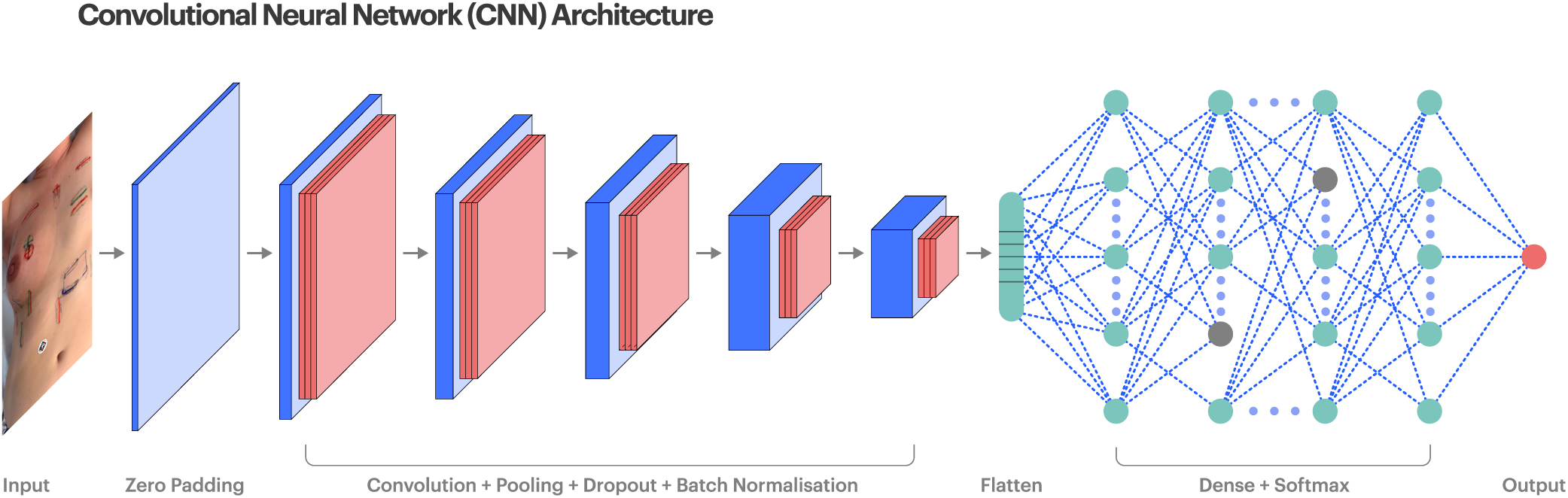
Convolutional neural network (CNN) architecture

At each node, the output can be computed as follows:

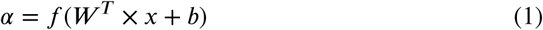

The softmax function is used as an activation function at the layer to convert the output values as probabilistic, which are added up to 1.

A neural network accepts an input as a vector, whereas a CNN can take both structured and unstructured data. CNNs have shown promising results in image classification and object detection [24, 25, 26]. The training process involves two steps: forward and backward propagation. A CNN algorithm consists of convolutional layers and subsampling layers, followed by a fully connected layer. The output from the convolutional layer can be computed as follows:

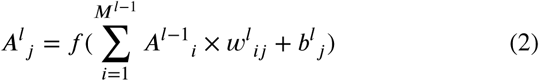

Where

*M*^*l*−1^ is the number of feature maps in the (l-1) layer,

*A*^*i*^_*j*_ is the activation output at the *l*^*th*^ layer,

*w*_*ij*_ is the kernel weights from feature map j at (l-1) layer, and *b*^*i*^_*j*_ is the bias.

The convolutional layers help in feature extraction, and maxpooling layers are mainly used for downsampling the features from convolutional layers, which is followed by a fully connected layer and consists of flattened and dense layers (with a ReLU and softmax activation layer). In convolutional layers, the filter weights shared by the CNN in each receptive field (particular layer) are the same, which ultimately reduces the memory storage of different weights and improves the performance of the CNN architecture. Convolutional layers also help in dimensionality reduction.

For this task, we addressed two main types of problems: For the first, binary classification is applied in which the CNN (Figure 5) was implemented to classify healthy (grade 0) versus erythema (grade ≥ 1) using the radiation oncologist-labeled images. For the second, a multi-class classification is applied, the task of which was to estimate the severity of the erythema into grades 0, 1, and 2 using a modified CNN. For a 2-class problem, the CNN algorithm consists of zero padding (ZeroPadding2D), which helps retain the relevant erythema features present at the corner or border points. The algorithm was followed by five convolutional layers (Conv2D). These and the pooling layers are used to retain the relevant features and reduce the dimensionality from padded input images, which particularly reduces the computation cost and improves the performance of the CNN model. Each convolutional layer was followed by a max pooling layer, batch normalization, dropout layers, the approach developed by Srivastava et al. [27], and and fully connected dense layers. The sigmoid activation layer was implemented to bring the output within the probabilistic range of 0 and 1, and a similar architecture was used for a 3-class problem.

We applied a dropout in convolutional layers because the dropout in an FCL is different. In an FCL, it is equivalent to zeroing out a column from the weight matrix associated with the FCL, which is basically applied to drop certain neurons from the neural network (to not train a neuron), whereas in a convolutional layer, this does not produce the same effect as zeroing out a column of the weight matrix corresponding to the convolutional kernel and still allows the weights in the column to be trained. We implemented a dropout in the convolutional layers to multiply Bernoulli noise into the feature maps of the network [28]. To prevent the model from overfitting (owing to a large number of total parameters and low input sample size, the model is prone to overfitting), we used dropout, early stopping, and reduced learning rate techniques while training the CNN. Normalization is conducted by subtracting the batch mean from each activation and dividing it by the batch standard deviation. Fine-tuning was applied to the following hyperparameters, including the number of output layers:

- Learning rate (α) = 0.0001
- Activation = relu, sigmoid, and softmax
- Optimizer = adam
- Batch_size = 32
- Padding = (3,3)
- Pool size = (3,3)
- Kernel_initializer = glorot_uniform
- Bias_initializer = normal
- Loss = binary_crossentropy, and categorial_crossentropy
- Metrics = accuracy
- Min (*a*) = 0.00001

#### 2.6.1. Ensembled convolutional neural networks (eCNN)

For classification tasks, ensemble learning was originally proposed in 1965; with the idea of training multiple base learners as ensemble members and combining their predictions into a single output, which should have a better performance on average than any other ensemble member with an uncorrelated error on the target datasets [29]. Many researchers have demonstrated the outstanding performance of ensemble learning for classification tasks [30, 31, 32, 33]. In ensemble learning, if one prediction is wrong, the other predictions will counteract this. The ensemble models are broadly categorized into ensemble models as follows:

- Majority voting or vote count
- Averaging
- Weighted average
- Stacking
- Blending
- Bagging
- Boosting

In this study, we implemented a combination of bagging and majority voting or vote count to create an ensemble learning model using a CNN. Bagging is mainly used when data must have variance; otherwise, bagging adds levels of additional iterations, and this approach irons out variance from a dataset. To verify the presence of variance in the input dataset, we trained and evaluated the CNN on multiple chunks (*L*\*, +*a*\*, +*b*\*, and *SEV* *) of the dataset separately and observed that the predictions were different, which confirmed such presence. In this experiment, the ensemble CNN consisted of two main approaches: first, bagging was used to split the dataset into different subsets to train and evaluate different iterations of the CNN, which was followed by majority voting to aggregate the prediction (maximum counts) to form a final prediction; in the case of an equal vote count, the prediction on the *SEV* * image was taken into account, resulting in a final prediction. In the paper “128 shades of Red”, we discussed about a higher dynamic application range of the measured erythema signal when compared to the signal parameter +*A*\* and the *SEV* * enables objective quantification of skin erythema. The *SEV* *, is also capable of measuring erythema based skin alterations by simultaneously providing a long dynamic range from bright to very dark skin tones (Fitzpatrick Type 0 to 5).

## 3. Results

### 3.1. Visual data exploration

From a visual assessment of the categorical variables, the total number of all scored grades 1, 2, and 3 was (n = 2263). To train the machine learning models, the statistical values (*SEV* *_mean, *L*\*_mean, +*a*\*_mean, and +*b*\*_mean) were computed by analyzing six regions of interest per image using the Scarletred^®^ Vision online platform. Most of the images came from breast cancer patients, whereas a few were from head and neck (HNC) or prostate cancer patients (Table 2). Here, *SEV* *_meandt-refdt shows a heavy tailed distribution for the areas scored with a CTCAE grade of 0, and similar results were observed for *L*\*_meandt-refdt (Supplementary Figure 14).

### 3.2. Comparison

Figure 6 shows boxplots for the measurements of all analyzed images separated by score. A clear trend in the *SEV* *_ meandt-refdt and +*a*\*_ meandt-refdt can be observed when the CTCAE grade from a visual assessment increases. By contrast, we can only observe small variations in +*b*\*_ meandtrefdt between the CTCAE grades. To see if the differences are significant, a Kruskal-Wallis test was conducted on a random subset of the data (n = 35 per group). The null hypothesis for this test states that there will be no significant difference for any of the groups. At a significance level of 5%, we can reject this hypothesis if the p-value is below 0.05. This is not the case for +*b*\*_ meandt-refdt (p-value of 0.0318) but is the case for all others (p value equal to or below 0.0001) indicating a highly significant difference between the groups in the latter parameters, which also supports the outcome of our previous study “128 Shades of Red” [16].

**Figure 6:**
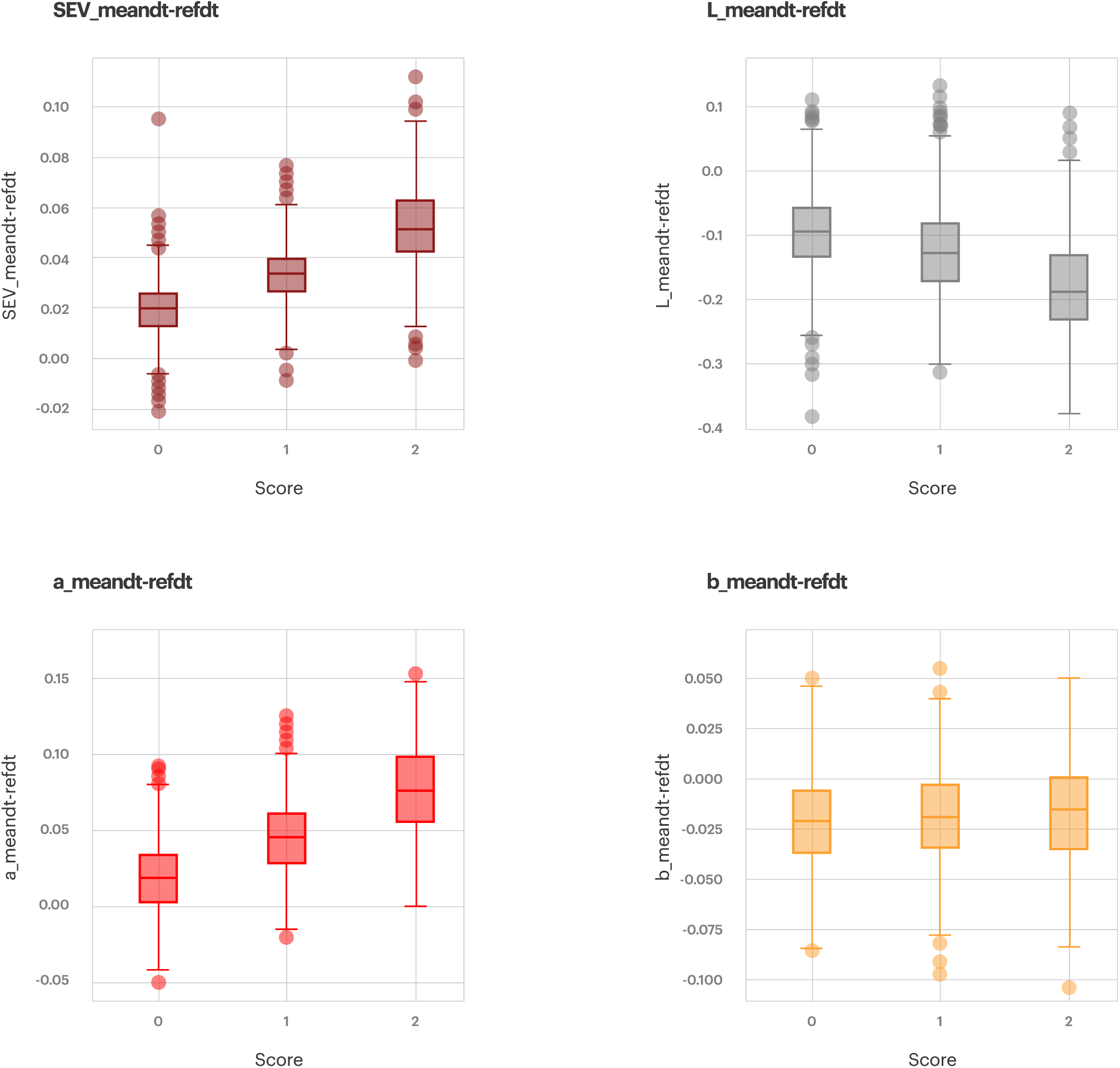
Boxplots for images with CTCAE-grades (score) of 0, 1 and 2 shown for every parameter (SEV *_meandt-refdt at the top left in dark red, L*_meandt-refdt at the top right in gray, +a*_meandt-refdt at the mid-point in light red, and +b*_meandt-refdt at the bottom in yellow).

### 3.3. Machine learning and CNN

Because of the promising results indicating a significant difference in the measurements between different CTCAE grades, different ML models have been tested regarding their capability to reproduce the CTCAE grading by using the measurement data. A support vector machine (SVM), decision tree, random forest and multinomial logistic regression (MLR) were tested. In parallel, a novel CNN model was developed to deduce the score directly from the pseudo grey images, representing the *L*\*, +*a*\*, +*b*\*, and *SEV* * signals.

#### 3.3.1. 2-class classification

First, the distinction between healthy (grade 0) and erythema (grade ≥ 1) was determined. After finding the best kernel for an SVM and feature list for all algorithms, the best model of each algorithm was used to compare their respective performances (Figure 7). All algorithms, including CNNs, achieved an accuracy of above 70%. The sensitivity of recognizing erythema was between 67% and 90% for all models. The specificity is the ability to classify only those patients with erythema, who also received such diagnosis through a visual assessment. Here, the specificity ranged from 72% to 83%. We can see that there are only slight differences in the performance metrics between the models with exception for the ensembled CNN, which clearly outperformed the all other algorithms as discussed later.

**Figure 7:**
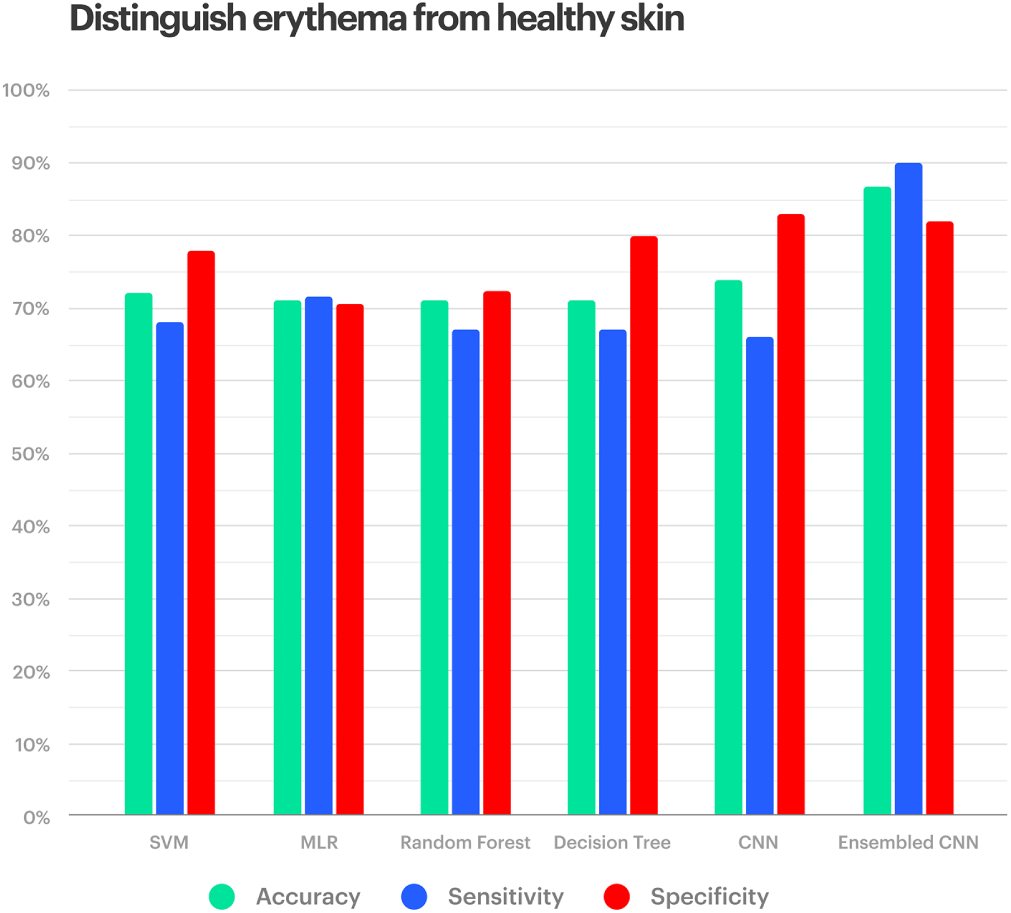
Different algorithms (from left to right: SVM, logistic regression, random forest, decision tree, CNN and ensembled CNN) have been tested for their performance in reproducing the visual assessment of a patient having erythema or not having erythema by using the measurements of the Scarletred^®^Vision system or the images themselves. Accuracy is represented in blue, sensitivity in green and specificity in red (left to right). On the y-axis the percentages of the results are given.

The good sensitivity and specificity of the SVM with the rbf kernel and *SEV* *_meandt-refdt as a single feature are also apparent in the corresponding confusion matrix of the model (Figure 8, upper left). A total of 78% of the healthy images were correctly classified, and 69% of the patients with erythema were also put into the actual category, when applying the SVM. The confusion matrix for the CNN model looks extremely similar with an 83% specificity and 67% sensitivity (Figure 8, lower left). The receiver operating characteristic (ROC) curves of both the machine learning and CNN models also showed a good performance (Figure 8, upper right). The curves for the classification between healthy and erythema are extremely steep with areas under the curves of 0.79 for machine learning, and 0.80 for the CNN, which are close to the maximal value of 1. Therefore, a high true-positive rate (sensitivity) is possible at a relatively low false-positive rate (high specificity).

**Figure 8:**
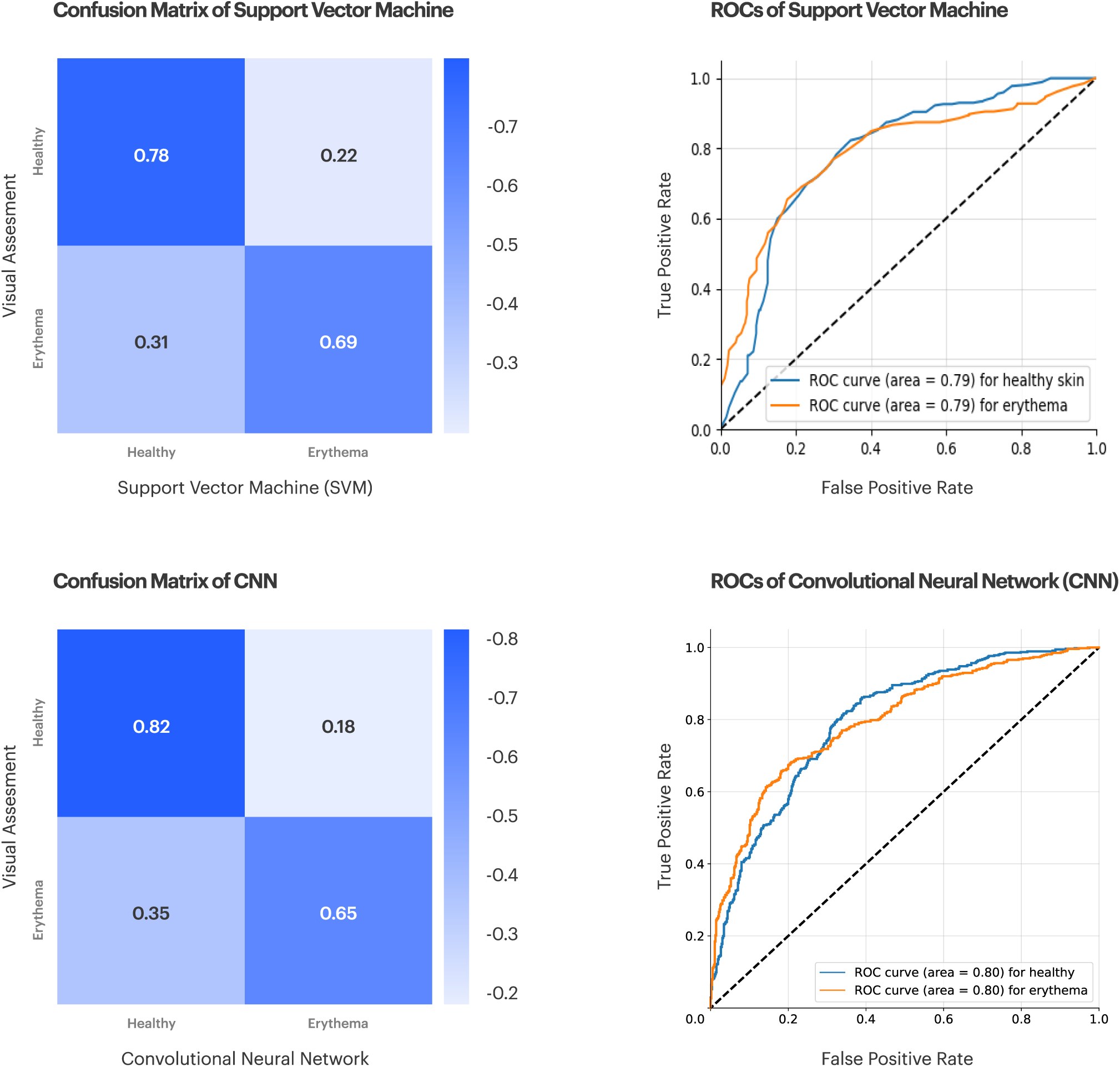
Confusion matrix and receiver operating characteristic (ROC) for the 2-class analysis using an SVM (upper row) and a CNN (lower row). The confusion matrix represents the score through a visual assessment and the SVM. The numbers in the squares represent the true negative rate or specificity (upper left), false positive rate (upper right), false negative rate (lower left), and true positive rate or sensitivity (lower right). The darker the square is, the higher the given rate. The receiver operating characteristic plots the false positive rate on the x-axis against the true positive rate on the y-axis for the classification as healthy skin (labeled 0 in blue) and the detection of erythema (labeled 1 in orange). The area under the curve is given in the legend (area). It should be close to 1, indicating a steep curve, which allows for a high true positive rate (sensitivity) at a low false positive rate (high specificity). The dotted line represents an area under the curve of 0.5 and represents the border, below which the classification can be seen as worse than a random guess.

#### 3.3.2 3-class classification

In the next step, we evaluate the performance of the different algorithms when distinguishing between all CTCAE grades (0, 1, 2). Again, the rbf kernel performs best with the SVM. The best results were achieved for all machine learning algorithms when using *SEV* *_refdt as a feature. Figure 9 shows the overall accuracy for the best model of every algorithm, as well as the sensitivity for every CTCAE grade. The overall accuracy was above 60% in all cases. The decision tree performs the worst in recognizing grade 1, and the best in recognizing grade 0. A logistic regression showed the highest specificity for every score. This model has two additional advantages over an SVM. On the one hand, it requires less computational power to build a logistic regression model. On the other, logistic regression provides formulas that make the results easy to follow and accept, whereas the SVM is more difficult to grasp. Therefore, we focused on the MLR to distinguish between different CTCAE grades.

**Figure 9:**
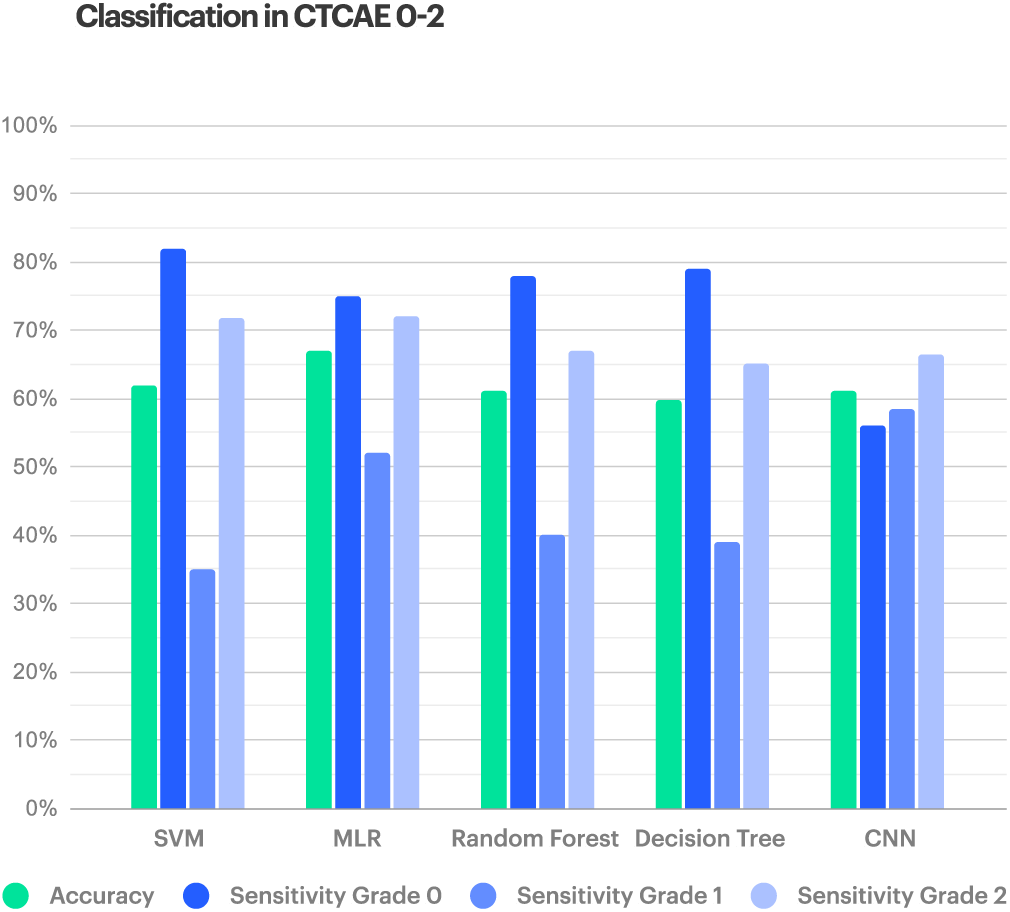
Different algorithms (from left to right: SVM, MLR, random forest, decision tree, and CNN) have been tested for their performance in reproducing the CTCAE-grades of the visual assessment.

The summary of the MLR model (Supplementary Figure 15) provides some background information and general estimators for this algorithm. We can see that it is built through a maximum likelihood estimation, converging against a loglikelihood of 594.58, which is significantly higher than, and therefore preferable to, the simpler null model (805.25). The pseudo R-squared, which represents the correlation between the scores from a visual assessment and the results from the model, is only 0.2616, which could be higher, because the maximal value is 1. The model assumes a CTCAE grade of zero. For each other CTCAE grade, a sub-model was built, resulting in a formula. The formula describes the logarithm of the odds (probability divided by the counter probability) as a result of the sum of the estimated intercept and each feature multiplied by an estimated coefficient. A hypothesis test was applied for each estimator. The null hypothesis states that the coefficient is not significantly different from zero. The null hypothesis can be rejected for the intercept and the coefficient of *SEV* *_meandt-refdt because the p-value is below 0.05. This makes those estimators relevant for our model.

For every increase in *SEV* *_meandt-refdt by one unit, the odds for a score of 1 is multiplied by 6.7 × 10^27^ (e to the power of the coefficient) and the odds for grade 2 is multiplied by 7.9 × 10^70^. From the odds the probability for grade 1 and grade 2 can be calculated. The counter probability of the sum of the probabilities for grade 1 and grade 2 is the probability for grade 0. The grade with the highest probability is chosen for every image. The confusion matrix (Figure 10, upper left) shows that the highest proportion of every grade from a visual assessment is classified as the same grade by multinomial logistic regression. This can be seen from the dark blue diagonal from the top left to the bottom right. A misclassification mostly occurred in grade 1. This is also the reason for the lower sensitivity in recognizing grade 1 because it can be misclassified in both directions, and all misclassifications from grades 0 and 2 were classified as grade 1. This explains the flatter ROC curve and area under the curve (Figure 10, upper right).

**Figure 10:**
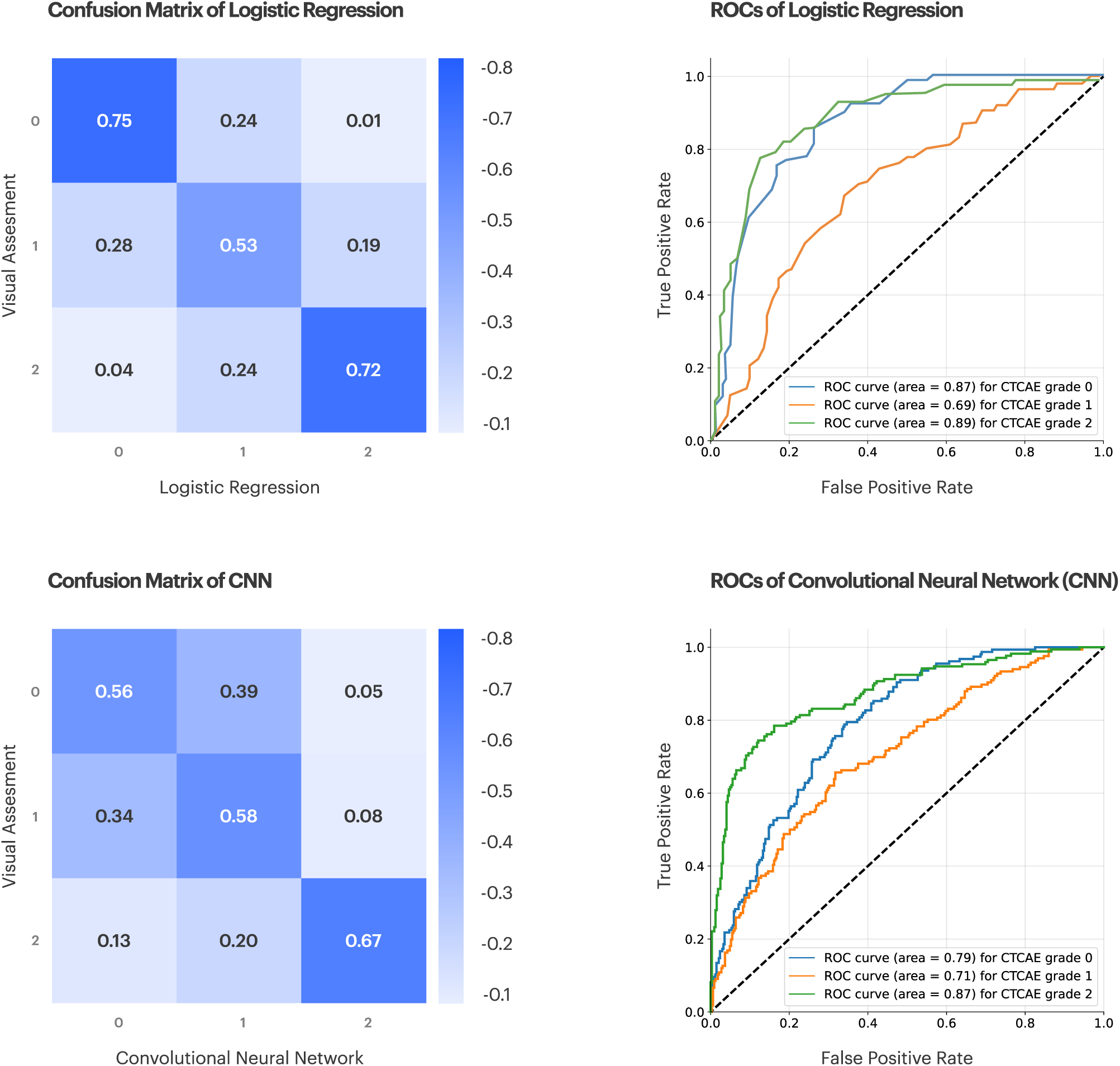
3-class confusion matrix and receiver operating characteristic (ROC) curves of the multinomial logistic regression (upper row) and CNN (lower row). The receiver operating characteristic plots the false positive rate on the x-axis against the true positive rate on the y-axis for the classification in different erythema grades (0 represented in blue, 1 represented in orange, and 2 represented in green).

The confusion matrix (Figure 10, lower left) shows the performance of the multiclass CNN classification algorithm for estimating the severity grade of erythema (0, 1, and 2). Ideally, in the case of multi-class classification, the perfect classifier would have the values only across the diagonal elements, which classifies all test samples correctly in the three classes. The values across the diagonal element are true positive (TP) values for a logistic regression, i.e., 75%, 53%, and 72% of the samples, were correctly classified, whereas for the CNN, 56%, 58%, and 67% were correctly classified as grade 0, grade 1, and grade 2, respectively, and a misclassification of grade 0 and grade 2 image samples as grade 1 also led to an increase in the false positive values.

The ROC curves for grades 0 and 2 for the CNN were as good as those in the previous classification problem. The less steep ROC curve and lower area under the curve for the classification in grade 1 by the CNN distinguishing the three grades of erythema (Figure 10, lower right) can also be explained. Therefore, it makes sense that in an unbiased model with equal sensitivity for all grades, and no misclassification over two grades, the middle grade will always receive less steep ROC curves with a lower area under the curve. The prediction accuracies were 73%, 66%, and 82%, sensitivities were 56%, 58%, and 66%, and specificities were 77%, 71%, and 93% for grade 0, 1 and 2, respectively.

#### 3.3.3 Ensembled CNN (eCNN)

In this study, a series of different machine learning models and an ensemble method were used to improve the overall prediction capability of the models. We evaluated the ensembled CNN (eCNN) on the same test dataset used for the CNN algorithm. The ensemble CNN consisted of bagging and majority voting techniques to form a final prediction; for a 2-class problem, the prediction accuracy of the ensemble model was 87% with a sensitivity and specificity score of 90% and 82%, respectively (Figure 7), while for each grade (0, 1, and 2) accuracies were 76%, 69%, and 87%, sensitivities were 70%, 57%, and 71%, and specificities were 78%, 75%, and 95%, respectively (Figure 11). Figures 12 and 13 show the confusion matrix for the ensemble model. Using an ensembled CNN for 2-class classification, 82% of the images from the healthy class were correctly classified, and the remaining 18% were incorrectly classified as erythema. Similarly, 90% of the samples from the erythema class were correctly classified, and the remaining 10% of the images were incorrectly classified as healthy. For a 3-class problem, the ensemble CNN was able to correctly classify 70% of grade 0 images, 57% of grade 1 images, and 71% of grade 2 images, as shown in Figure 13.

**Figure 11:**
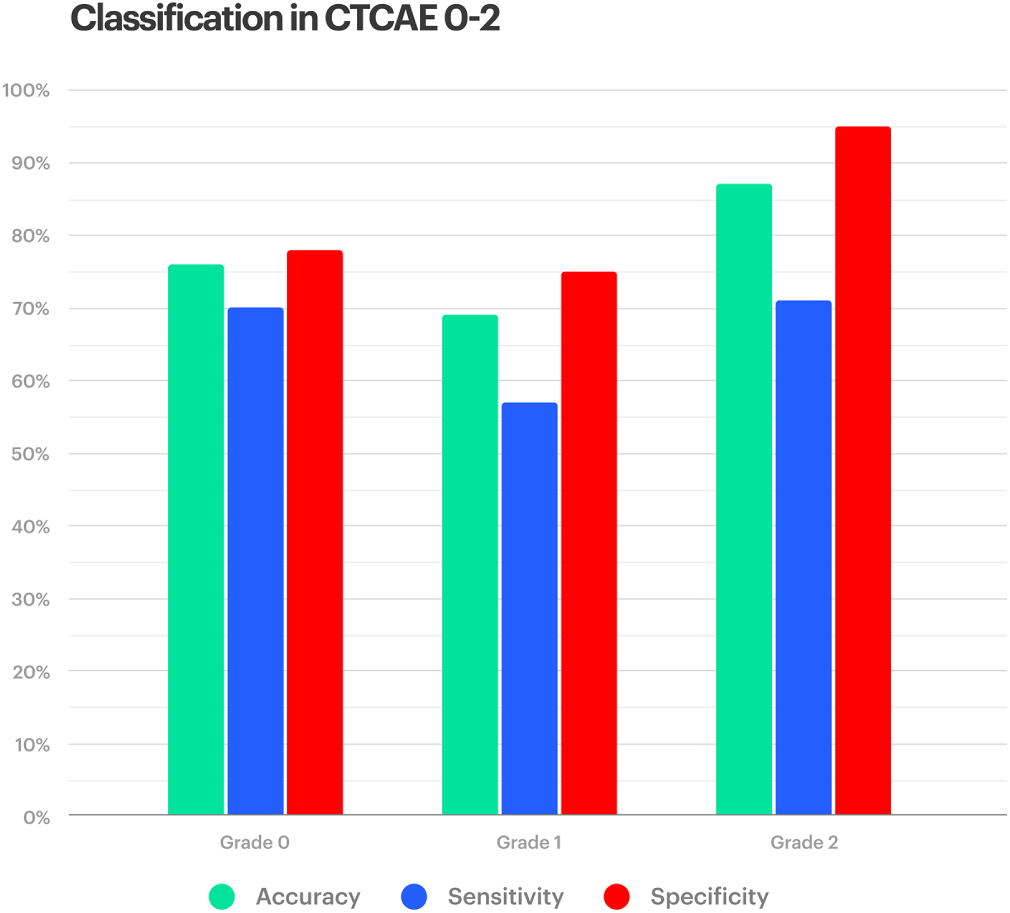
Class-wise performance of ensembled CNN: Prediction accuracy, sensitivity, and specificity for CTCAE 0-2

**Figure 12:**
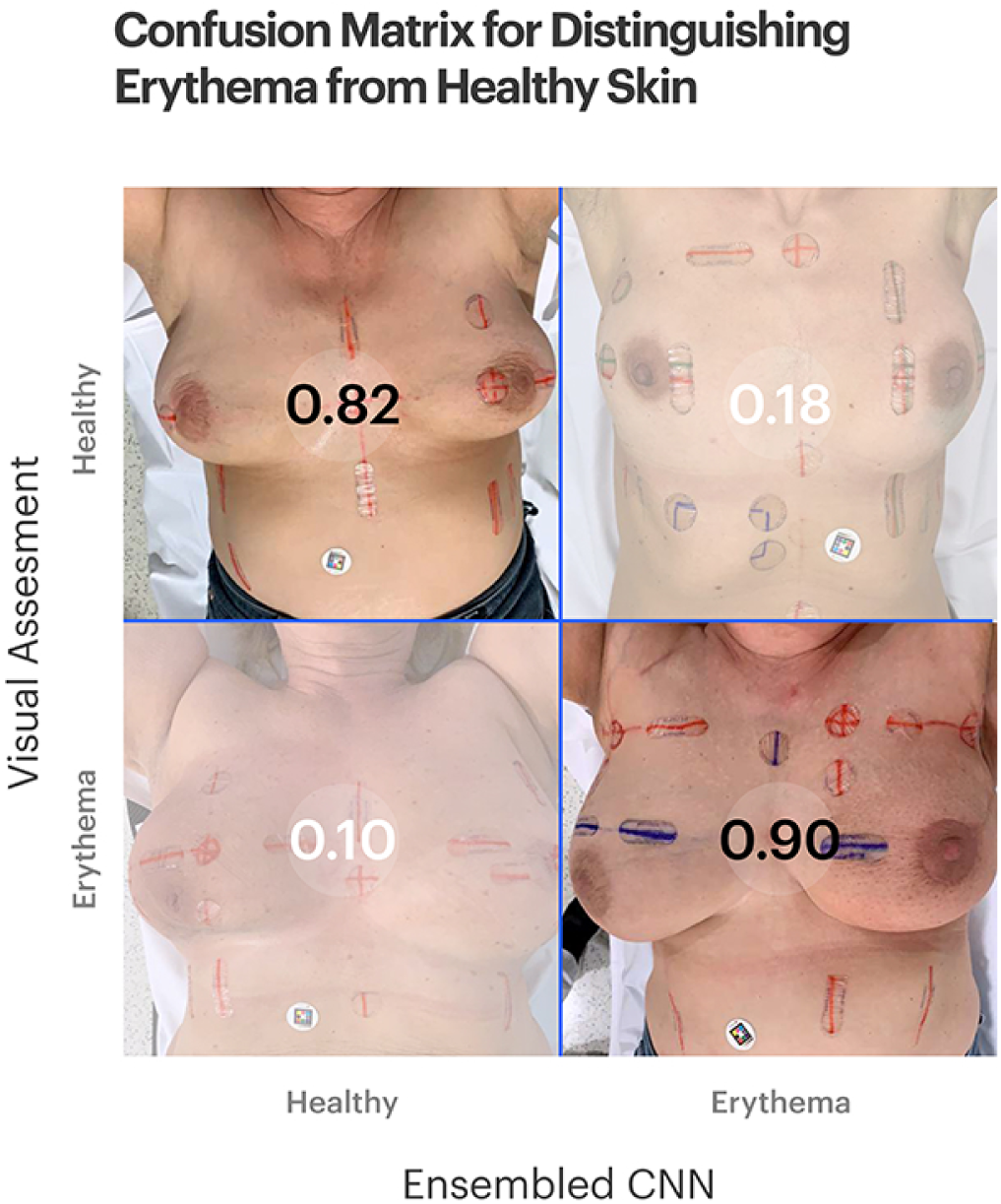
Randomly selected examples of correct and incorrect predictions for healthy (grade 0) versus erythema (grade ≥ 1) classification, displayed as a 2×2 confusion matrix. The upper-left quadrant shows an example of true negatives (n_00_ = 99 images), whereas the lower-right quadrant shows an example of true positives (n_11_ = 157 images). A false-positive (n_01_ = 22 images) is given in the upper-right quadrant, and a false negative (n_10_ = 17 images) in the lower-left quadrant

**Figure 13:**
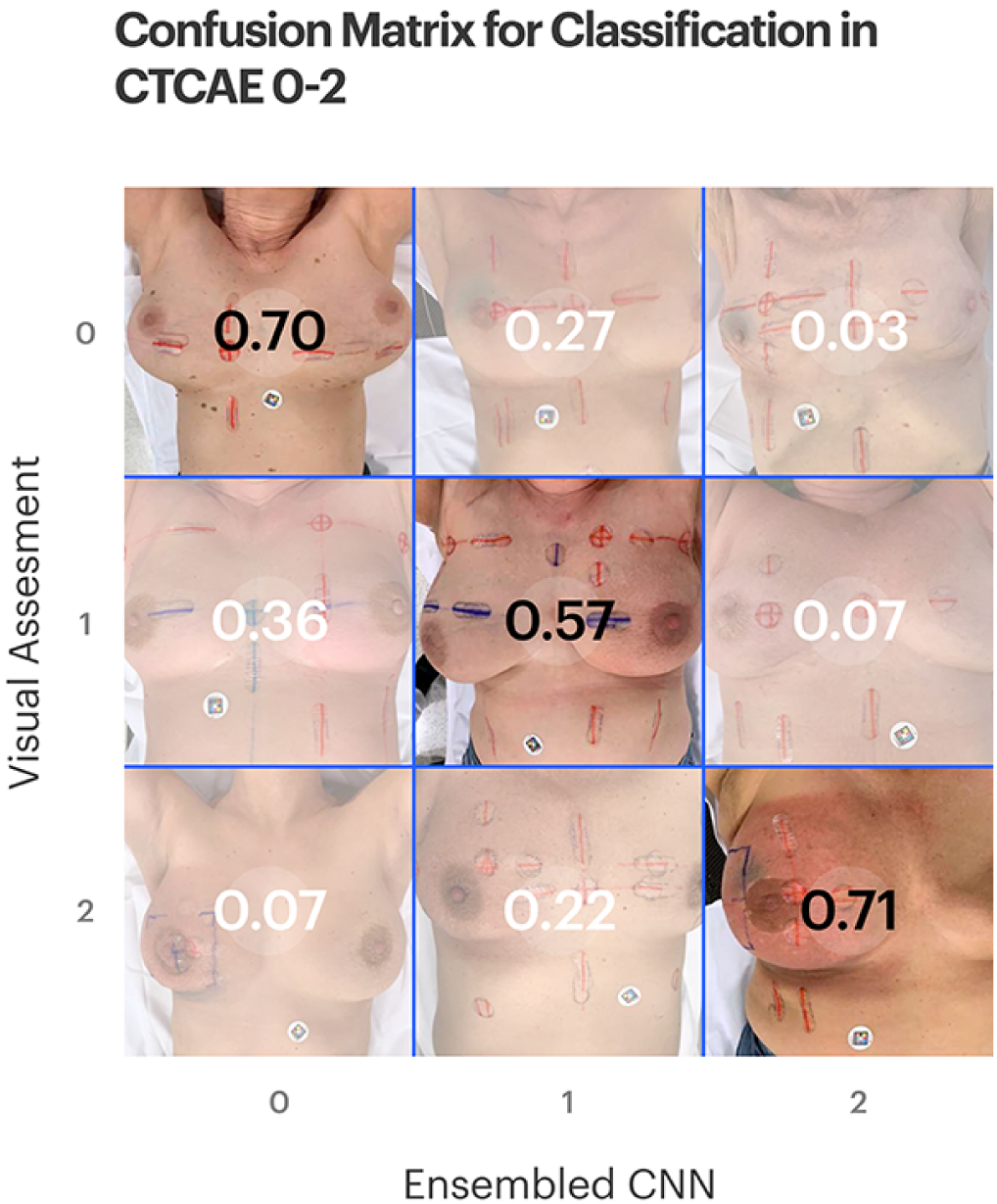
Randomly selected examples of correct and incorrect predictions for classification in CTCAE grade 0 versus grade 1 versus grade 2, displayed as a 3 × 3 confusion matrix. The diagonal elements are the true positive values. The perfect classifier would have elements across the diagonal, which means classifying all test samples correctly in three classes.

## 4. Discussion

The high demand for objective scoring in dermatology, supported by a trend toward digitization, confronts medical experts and clinical research organizations with a large variety of newly developed methods. However, in addition to the requirement that a new method needs to build upon internationally established standards, ideally it should be easy to use and require only little or no specialized equipment in order to increase acceptance and enable fast technology adoption in clinical routine or trial procedures. The digital solution presented in this paper automates the assessment of skin erythema and RISRs. It only requires a smartphone with an installed Scarletred^®^ Vision mobile app, the corresponding calibration sticker provided by SCARLETRED, and a computer to obtain access to our secure web platform. In addition to the benefit of requiring only little equipment, data management and analysis is fully automated by the CNN, which enables the medical expert to save valuable time in a clinical routine and increases the learning curve in clinical research.

In a different study by another group, an automated technique [34] was proposed to classify nine types of skin cancer. In addition, Rezaoana et al. proposed an image processing and convolution neural network approach with image augmentation techniques. They used a transfer learning approach to implement the CNN, whereas in our case, we specifically focused on solving the problem of classifying RISRs and estimating the severity of skin erythema on the basis of calibrated skin images. Given an enriched number of inputs in the dataset, their study reported an accuracy of 79.45%. Melanoma skin cancer is one of the most lethal diseases, and in 2016, automated melanoma recognition by leveraging an extremely deep convolutional neural network (deep-CNN) with more than 50 layers [35] was proposed for specific lesion categorization. Hao et al. proposed the separation of lesion areas from the input images, and a deep residual network (DRN) was used to identify melanoma and non-melanoma lesions with a test accuracy of 69.3%. This framework was evaluated on the ISBI 2016 Skin Lesion Analysis Towards Melanoma Detection Challenge dataset.

In 2015, however, for medical image segmentation and pathology in computer modalities, the U-Net was proposed, which was effectively used in the medical domain, particularly for image segmentation, and obtains a reliable performance [36]. Later, a recurrent residual convolutional neural network U-Net (R2U-Net) was introduced, which produced a significant improvement using the ISIC 2017 dataset for skin cancer image classification tasks [37]. This same year, a U-Net-based segmentation approach using the concept of the triple attention mechanism was also proposed [38], and in this study, Tong et al. first selected regions using an attention coefficient computed using the attention gate and contextual information. Second, spatial attention and channel attention were used as a dual attention decoding module to capture the spatial correlation features and improve the skin lesion segmentation performance by applying the ISIC-2016, ISIC-2017, and PH2 datasets. This triple attention mechanism helped the network focus on a more relevant field of view of the target.

Dermatologists are dealing with a multitude of different cutaneous disorders in their daily work, and artificial intelligence can thus be harnessed to reduce the risk of potential errors by providing a fast decision support on the severity classification of a specific disease and in the diagnosis of the disease itself. As an example, to address the challenges associated with detecting erythema migrans (EM), i.e., a rash of acute lyme disease, versus multiple other skin conditions, including cellulitis, tinea corporis, herpes zoster, erythema multiforme, lesions from tick bites and insect bites, and non-pathogenic normal skin, Burlina et al. [39] proposed a deep neural network that utilizes different pre-trained models such as ResNet50, InceptionV3, Mobile NetV2, DenseNet121, Inception Res NetV2, and Res Net152. The test accuracy ranged from 71.58% for an 8-class problem of EM versus seven other classes to 94.23% for a binary problem of EM versus non-pathological/healthy skin.

Raina et al. [40] proposed a similar scoring method in 2015, using a camera and a color card to obtain standardized color images. They used a wide variety of parameters to reproduce the scores from the visual assessment using a linear discriminant analysis. The general accuracy of the prediction was 48.75% when distinguishing between the five grades. However, it should be noted that linear discriminant analysis places some demands on the data distribution of the data it is used upon. When these demands are not fulfilled, the classification may worsen drastically [41]. This is not the case with our developed methods. Therefore, the proposed method is more robust. In addition, our method uses fewer parameters, which makes it simpler, more understandable, and easier to integrate in routine research. However, our study focuses exclusively on erythema in radiation dermatitis, a single skin parameter that is of central importance in the scoring of the majority of skin diseases, such as psoriasis or atopic dermatitis, and our erythema scoring method needs to be supplemented by additional parameters such as induration, desquamation, the affected body part, and the area in percentage.

Misclassification of the machine learning models may be explained by the fact that the measurements in the different grades of erythema lie close together. Figure 6 shows that the median 50% of the values for *SEV* *-meandt-refdt do not overlap. However, the outer 50% of the datapoints (whiskers), and those datapoints classified by the plot as outliers, do show a major overlap. If the same measurement can be found in two or even all three classes, it is understandable that the algorithms will face certain challenges. This also explains why grade 1, which is the middle grade, is particularly difficult to recognize for all algorithms. The measurements of this score can always be found in the higher or lower grade as well, although it may have a lower probability. The reason for the overlap might be that the visual assessment may also be based on factors other than the measured redness (e.g., dryness or scaling). In the first three grades, the main factor was redness. The other explanation for the overlap would be the subjectivity of a visual assessment, classifying the same intensity of redness as grade 0 or grade 1 in different patients or under different settings. This question needs to be further investigated. The machine learning models may improve when taking the visual assessment from multiple physicians into account because the opinion of a group of physicians (multimodal score) is assumed to be less subjective than the opinion of a single physician.

The classification of skin images through a CNN has attracted the attention of researchers, dermatologists, and computer scientists because of its potential to increase the intelligibility of skin cancer screening and streamline the workflow of dermatologists and radiation oncologists. Deep learning algorithms have recently shown promising results in visual tasks, such as ophthalmology, pathology, and radiology [42]. According to the paper published by Brinkler et al., the CNN can exhibit a superior sensitivity and specificity in melanoma classification as compared to board-certified dermatologists [43]. CNNs have already found their way into many aspects of our lives, including image analysis. When applied to a medical image analysis, there is still some hesitation in most people to consider the results. Although the concepts of a logistic regression and random forest models can be explained, a CNN is still perceived as a black box where data go in and a result comes out, without knowing exactly what happens inside. In response, no one wants to take responsibility for errors that might be caused by this mysterious method [44].

Therefore, we strive to improve the acceptance of a CNN in medical applications by showing that it is equally as trustworthy as many of the more comprehensible methods. While evaluating and understanding the results of the CNN classifier, it is important to note that accuracy, sensitivity, or specificity might not provide the exact performance of the classifier model, which is why the ROC is also reported with other performance metrics in our study. A classifier should not be judged merely on the basis of specificity, sensitivity, and accuracy [45]. According to Han’s response, the true ability of the classifier is indicated by the ROC curve to perform under a wide range of thresholds during classification using the input images. In this study, we used a radiation oncologist labeled image dataset to train a set of different machine learning and deep-CNN based algorithms that use image labels and raw pixel values for automatic classification of RISRs and the severity grading of erythema.

The machine learning and deep-CNN based algorithms were validated by comparing the severity grades of erythema given by the algorithms against those given by radiation oncologists. We found that the CNN algorithm helps in by-passing all segmentation and handcrafted feature extraction approaches. However, normalization can be introduced to improve the prediction ability of the CNN algorithm. The reasons behind certain misclassifications could be due to the fact that, for each image, the entire part of the body from head-neck to pelvis was considered for training the classification algorithm instead of only the erythema region. Such misclassifications might also be due to the presence of ink marks, which certainly act as additional noise to the CNN, which in some cases may be overcome by introducing adversarial networks; in addition, the region of skin under the ink marks may also be reconstructed to improve the performance of the CNN algorithms. Moreover, different image augmentation techniques such as histogram equalization or noise filtering can be introduced to reduce the changes in misclassification or bias from the algorithm because the intensity or pixel distribution is uneven in the input images, which may act as noise (or provide bias) to the CNN algorithm. Thus, our future work may include using a larger set of datasets to reduce the changes of an overfitting, improved regularization, fine-tuning of the hyper-parameters, and implementing transfer learning approaches. In recent years, there has been a large increase in the use of transfer learning algorithms such as AlexNet, InceptionV3, and VGGNet, in different ways, such as: (1) training the pre-trained algorithm from scratch, (2) transfer learning from a pre-trained model (ImageNet), and (3) transfer learning and fine-tuning the CNN algorithm. We may further include approaches that combine a deep CNN with Fisher vector encoding and an SVM classifier [46]. Instead of using the entire image as input to the CNN, samples or sub-samples may be given to the CNN as input to reduce the chances of an over-fitting.

Further studies may also appear if the error rate of any of these algorithms may be comparable to the error rate occurring when comparing scores of the same subject from different physicians owing to the subjectivity of the visual assessment. In this case, the results from the ML algorithms would have the advantage of being reproducible, even though they may differ from a subjective visual assessment. By comparing the performance and other advantages and disadvantages of our CNN model with more comprehensible models such as a logistic regression, where the results can be brought back to mathematical formulas, our goal is to increase the acceptance of both methods as a decision support system. Such a system is due to its inherent lack of guaranteed correctness, and was not made to replace, but merely support, the visual assessment of a physician. However, the direct interaction between man and machine should be enabled in the future to avoid major misclassifications either through a visual assessment or through a computer-aided decision support system.

As one of the main limitations of this study, mainly Caucasians, and thus patients with lighter skin tones (Fitzpatrick scale 1–3) were included. Its applicability to populations outside this skin type is uncertain. Currently, this study is focused exclusively on erythema in radiation dermatitis, and the first benchmark results were produced using the ML and deep CNN models. Further refinements in future studies with greater skin diversity are warranted. Later, this study can be extended to analyze multiple skin parameters and/or disorders within a single image. With a higher diversity and greater quantity of data, including signal intensity mapped images along with depth and texture images, the results of this study may lead to additional improvements, thereby opening up a broad range of novel AI-supported applications in the field of digital dermatology and teledermatology.

## 5. Conclusions

A set of different AI-based algorithms was explored in this study for the automatic classification of RISR images. Based on our results, we demonstrated that our currently used ML and deep learning algorithms can serve as a useful decision support in scoring the erythema severity and automatic pre-screening of skin toxicity in individual patients. Furthermore, this method may identify patients who need intensive local skin care during or after cancer irradiation treatment.With the assistance of our AI based methods, physicians can save time and efficiently reduce the workload in their daily routine by easily integrating the novel state-of-the-art AI-based erythema scoring method in the form of a decision support system during their clinical trials.

## Data Availability

Since the raw data (skin images) relates to sensitive health data from cancer patients, therefore the data cannot be made available under the public domain.

## 6. Ethical conduct of the study

This clinical study was carried out under the title “Validating the method for objective remote assessment of radiation dermatitis by augmented digital skin imaging using Scarletred^®^ Vision”, and approved under the Ethics Committee (EC) number 30-023 ex 17/18 on the date of 06. April 2018, by the EC board of the Medical University of Graz, Augenbruggerplatz 2, A-8036, Austria. The study has been carried out in accord with the Declaration of Helsinki and all applicable laws and regulations of Austria, where the study was conducted, and in compliance with the current Good Clinical Practice guidelines (CPMP/ICH/135/95).

## Supplementary materials

**Figure 14:**
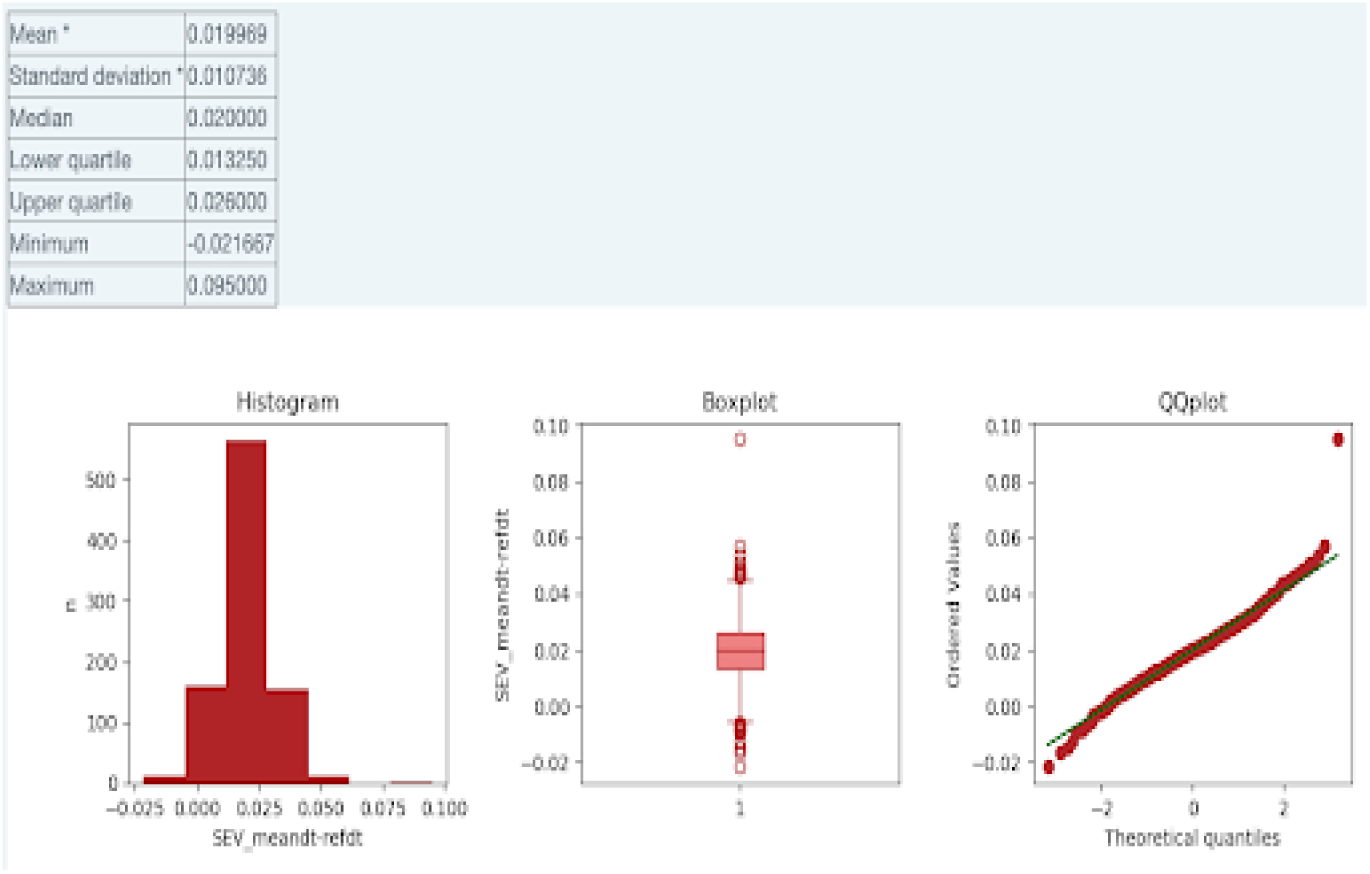
Output of the biostatistics tool for the parameter SEV*_meandt-refdt for areas scored with a CTCAE of zero. The table of estimators includes information on the mean, standard deviation, median, lower- and upper quartile, minimum, and maximum. This histogram plots the values of SEV*_meandtrefdt on the x-axis against the number of images within a certain value range on the y-axis. The boxplot shows the values of the parameter on the y-axis. The box ranges from the lower to the upper quartiles. The vertical line in the middle of the box indicates the median. The whiskers reach the minimum and maximum, respectively, excluding outliers. Outliers are datapoints that are more than 1.5-times the inter-quartile range away from the upper or lower quartile and are represented as small circles. The QQ-plot plots the ordered values of the parameters on the y-axis against the quantiles of a normal distribution on the x-axis.

**Figure 15:**
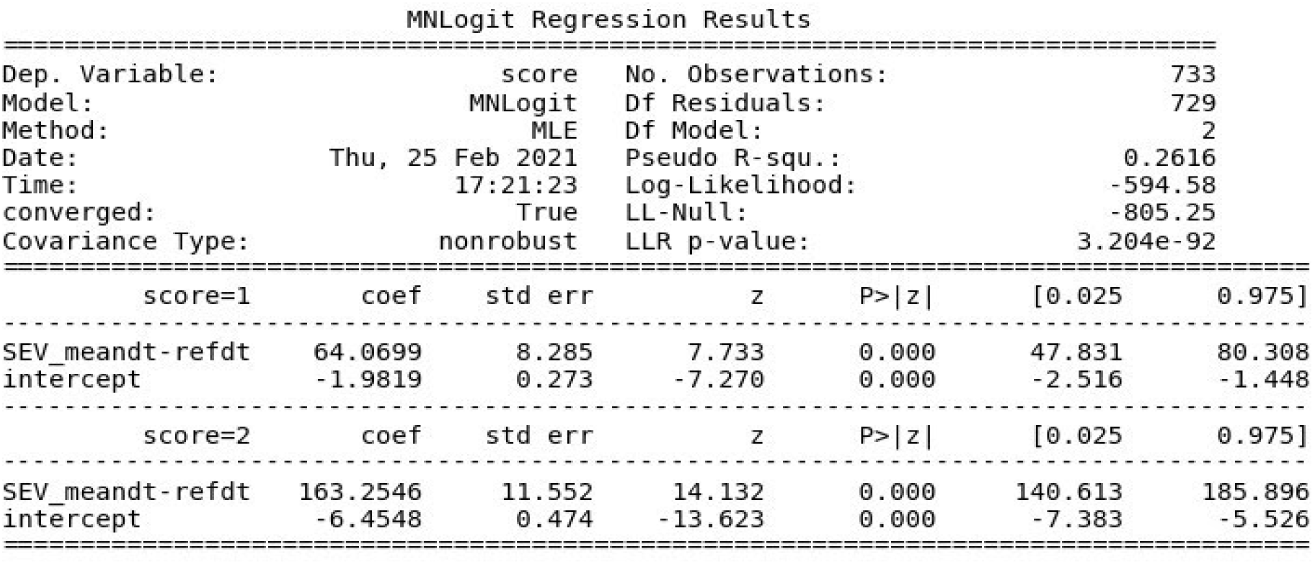
General parameters of the multinomial logistic regression model (MNLogit) explaining the dependent variable (Dep. Variable). The model was built using a maximum likelihood estimation (MLE), converging against the maximal log-likelihood. In any case, this value should be higher than the maximal likelihood of the null model, excluding only the intercept (LL-Null). In general, an LLR p-value of below 0.05 means that we can reject the null hypothesis that a more restricted model would yield significantly better results. The degrees of freedom for the residuals (DF Residuals) are calculated from the number of observations (No. Observations) minus the number of estimated coefficients. The degrees of freedom for the model (Df Model) are calculated from the number of estimated coefficients minus the number of submodels. Pseudo R squared (Pseudo R-squ.) should be as close to 1 as possible, indicating a high correlation between the real dependent variable and that estimated by the model. The model generally assumes a score of zero. The second part of the summary describes a submodel for each score, which differs from zero. For each submodel, the estimated coefficients (coef) for each feature and the intercept are given, including the standard error (std err) and z-score (z) as well as the 0.025 and 0.975 quartiles of the estimation. If the p-value (P>|z|) is below 0.05 the null hypothesis that the coefficient is not significantly different from zero can be rejected at a significance level of 0.05, and thus the feature is relevant for the submodel. Otherwise, the feature can be eliminated from the list of variables used to predict the score in the settings. The multinomial logistic regression algorithm assumes a CTCAE grade (score) of zero. It generates formulas that describe the logarithm of the odds (probability against counter-probability) for the classification under another specific CTCAE grade, using the provided features.

**Figure 16:**
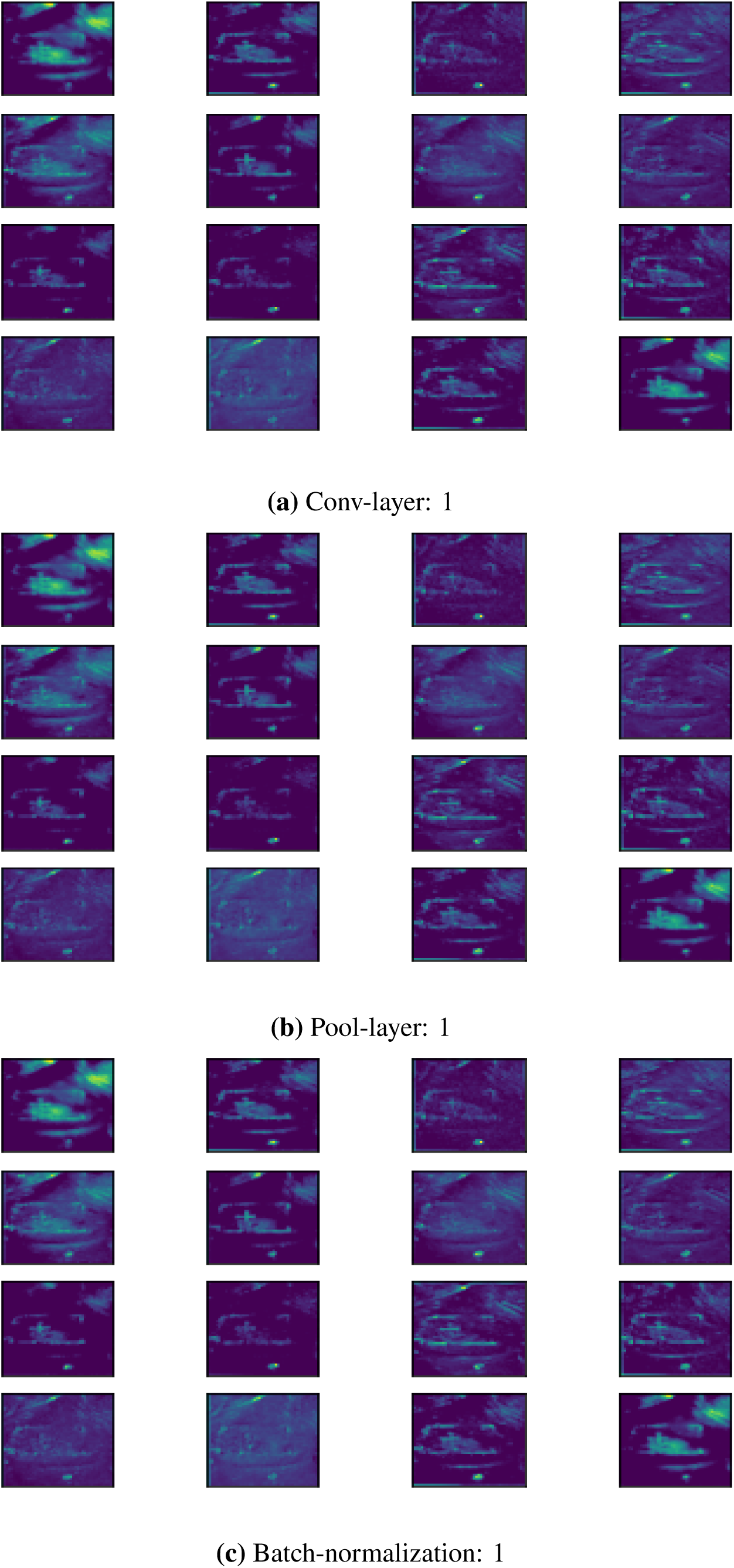
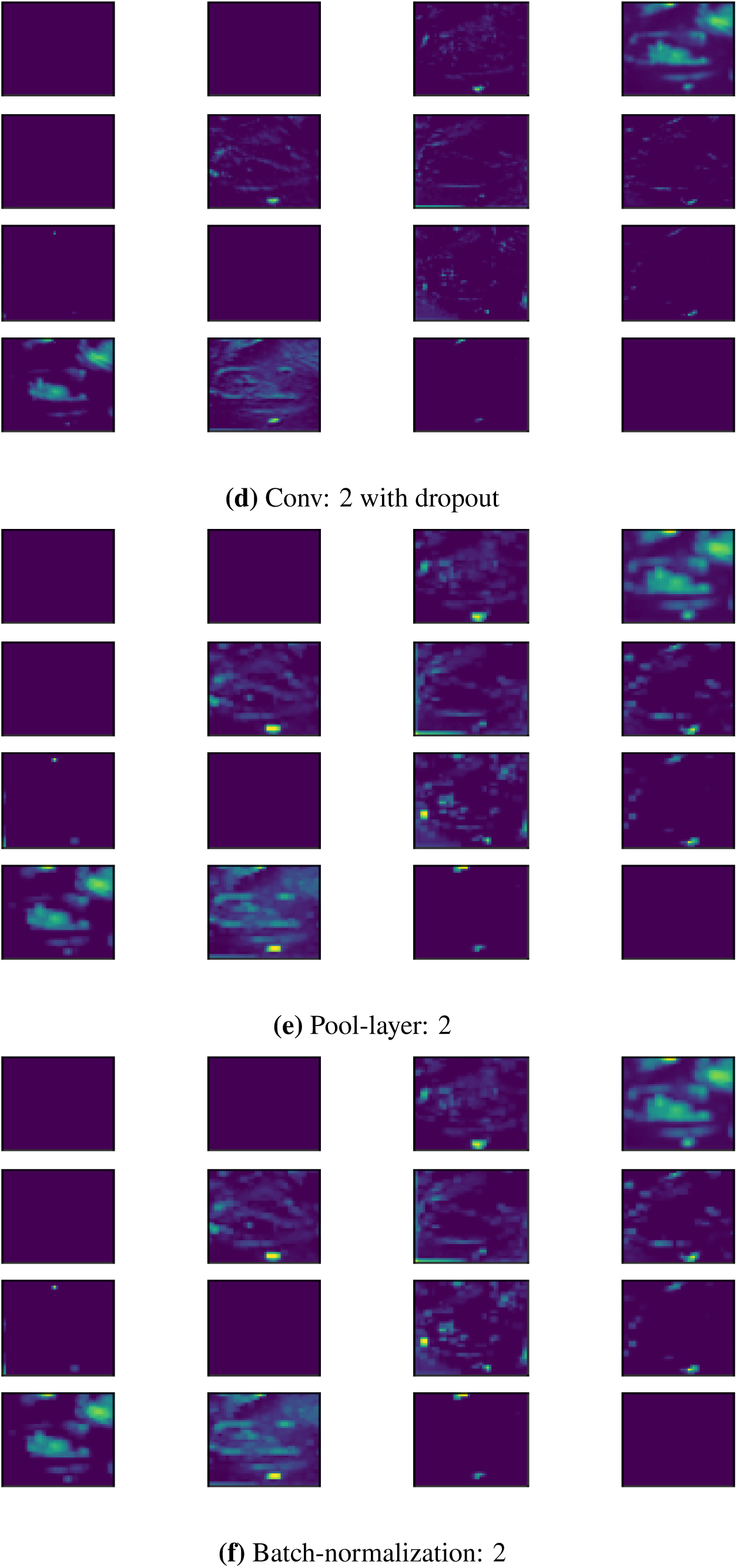
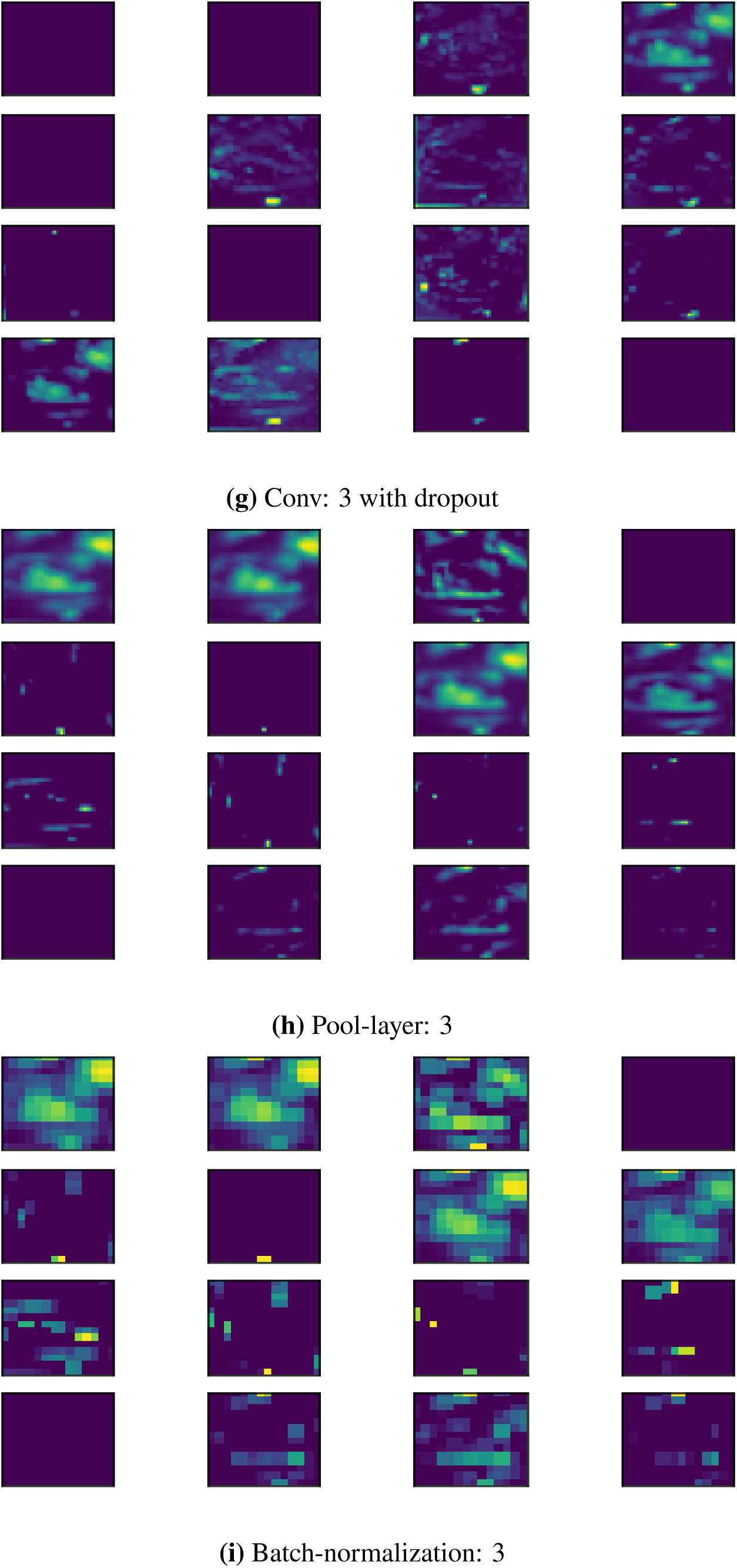
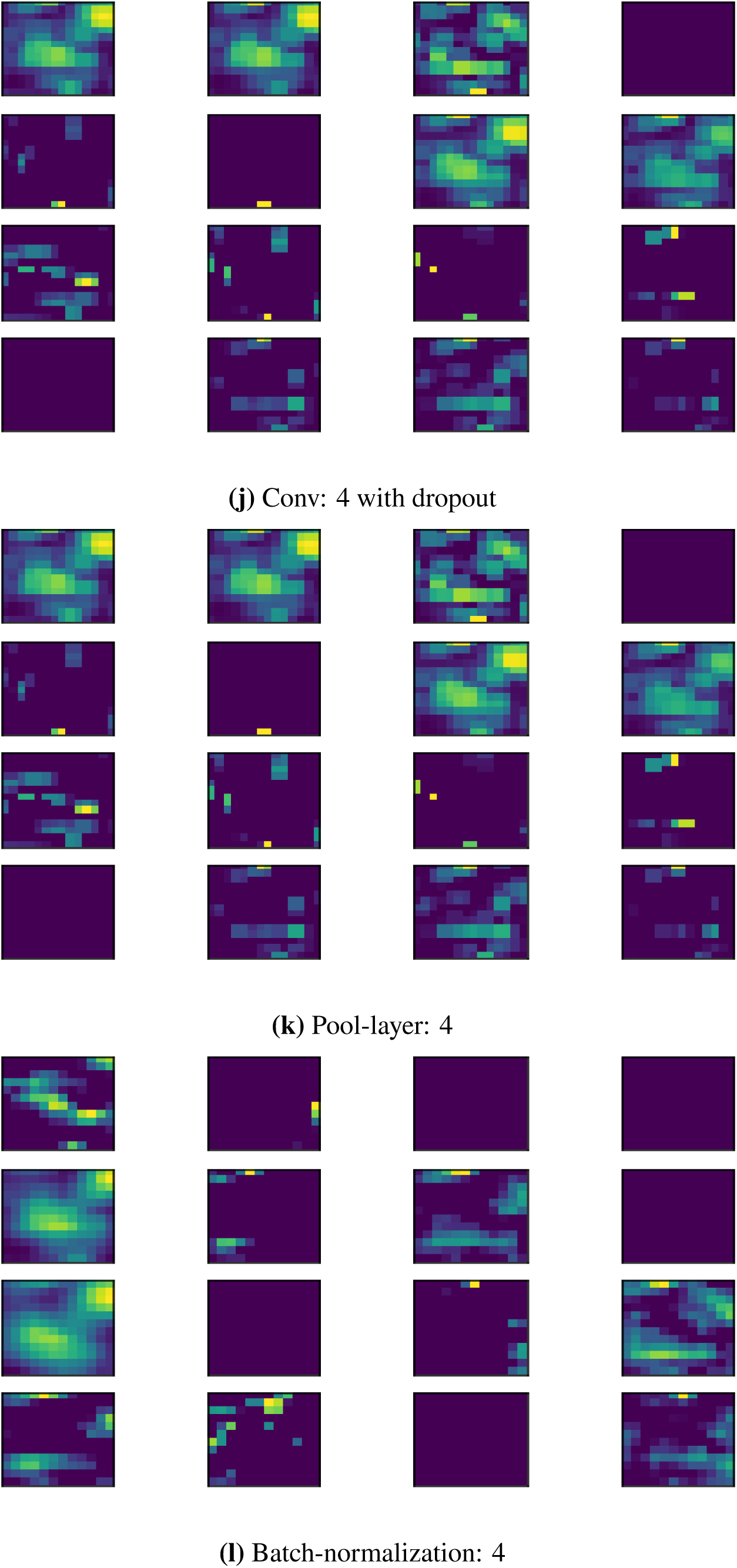
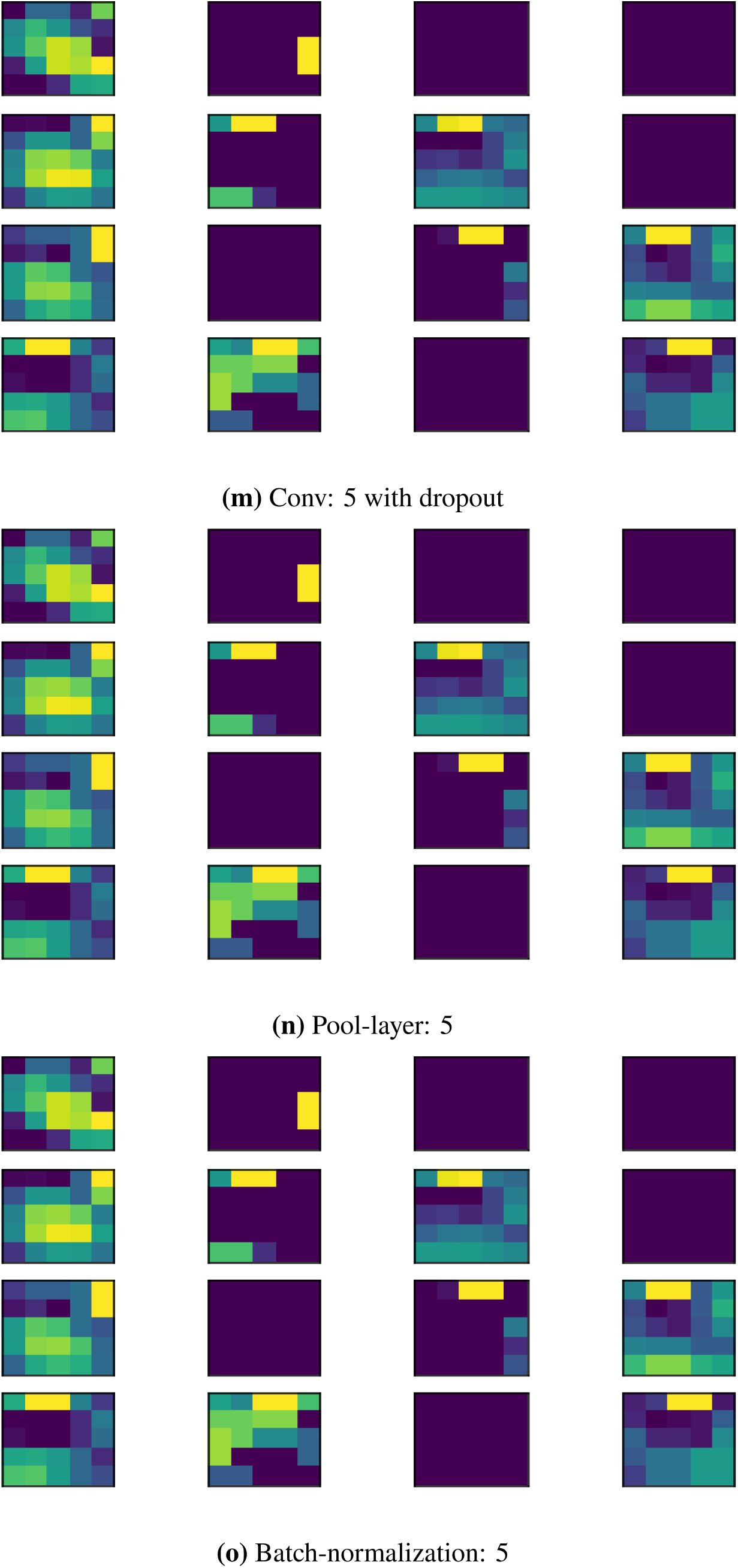
Visualization of convolutional feature maps

